# Interplay of global multi-scale human mobility, social distancing, government interventions, and COVID-19 dynamics

**DOI:** 10.1101/2020.06.05.20123760

**Authors:** Aniruddha Adiga, Lijing Wang, Adam Sadilek, Ashish Tendulkar, Srinivasan Venkatramanan, Anil Vullikanti, Gaurav Aggarwal, Alok Talekar, Xue Ben, Jiangzhuo Chen, Bryan Lewis, Samarth Swarup, Milind Tambe, Madhav Marathe

## Abstract

This work quantifies mobility changes observed during the different phases of the pandemic world-wide at multiple resolutions – county, state, country – using an anonymized aggregate mobility map that captures population flows between geographic cells of size 5 km^2^. As we overlay the global mobility map with epidemic incidence curves and dates of government interventions, we observe that as case counts rose, mobility fell and has since then seen a slow but steady increase in flows. Further, in order to understand mixing within a region, we propose a new metric to quantify the effect of social distancing on the basis of mobility.Taking two very different countries sampled from the global spectrum, We analyze in detail the mobility patterns of the United States (US) and India. We then carry out a counterfactual analysis of delaying the lockdown and show that a one week delay would have doubled the reported number of cases in the US and India. Finally, we quantify the effect of college students returning back to school for the fall semester on COVID-19 dynamics in the surrounding community. We employ the data from a recent university outbreak (reported on August 16, 2020) to infer possible R_eff_ values and mobility flows combined with daily prevalence data and census data to obtain an estimate of new cases that might arrive on a college campus. We find that maintaining social distancing at existing levels would be effective in mitigating the extra seeding of cases. However, potential behavioral change and increased social interaction amongst students (30% increase in R_eff_) along with extra seeding can increase the number of cases by 20% over a period of one month in the encompassing county. To our knowledge, this work is the first to model in near real-time, the interplay of human mobility, epidemic dynamics and public policies across multiple spatial resolutions and at a global scale.

## 1 Introduction

The COVID-19 pandemic is arguably the most acute public health emergency since the 1918 influenza pandemic. It has already infected over 16 million people and resulted in 646K deaths across the globe^1^. The economic impact is expected to be 3-10 trillion dollars^2^. The pandemic has affected almost every country in the world^3^ and has resulted in an unprecedented response by governments across the world to control its spread. Pharmaceutical interventions are not generally available at this stage (with the exception of remdesivir under FDA’s expanded access [1]) and thus, countries have had to rely exclusively on behavioral interventions that involve some form of social distancing. The rapid spread of the pandemic has forced countries to institute strict social distancing measures. Each social distancing policy is characterized by: (*i*) when and how gradually it started, (ii) the length of time for which it was enforced, (*iii*) the scope and pervasiveness, and (*iv*) the stringency (total lockdown versus stay-at-home advisories). For example, across the US, social distancing policies were instituted at the state level in a progressive manner: a declaration of a state of emergency by many states, followed by school closures, with the final extreme measure being stay-at-home or shelter-at-home orders (SAHO) [2], which saw the closure of recreational centers, parks, restaurant dine-in services, etc. As a contrast, a country-wide lockdown was instituted in India which subsequently led to suspended road, rail, and air transport. Non-essential services and schools were closed and individual-level mobility was severely curtailed.

Evaluating the level of public response to the global, country, and state level restrictions is important to understand COVID-19 dynamics. However, it is important to do this without compromising individual privacy. As pointed out by multiple public health experts and demonstrated in the literature [3, 4], aggregate mobility data, acquired through mobile phone location history, global positioning systems data, direction requests data, etc., indicate a considerable reduction of activity by individuals during the pandemic, and thus act as surrogate data sources to understand compliance with social distancing measures. With appropriate data-sharing policies, these data sources can be used to study social distancing, while also ensuring individual privacy through a combination of anonymization, aggregation and noising techniques that provide the needed privacy guarantees. There have been a number of recent studies along these lines, for example, in China using Baidu data, in the US using mobility data, and at a global scale using airline traffic [5, 6, 7, 8, 9, 10, 11, 12, 13, 14, 15]

The Google COVID-19 Aggregated Mobility Research Dataset (cf. Appendix A, henceforth called interchangeably mobility map or mobility flows (MF)) provides a global, time-varying anonymized mobility map of flows at a resolution of 5km^2^. Figure 1 provides an overview of the volume of local mobility at the level of cell resolution, county, state, and country. The data set has guaranteed differential privacy while capturing mobility flows (MF) at every level of spatio-temporal resolution. The global coverage enables us to undertake multi-scale global analyses of changes in mobility patterns. We combine this data with two additional data layers: data from the University of Virginia COVID-19 Dashboard^4^ that provides detailed, global epidemic surveillance data and data on social distancing guidelines in US states and India (at the national level). The integrated layered map provides a new way to assess the impact of changes in global mobility patterns on COVID-19 dynamics. Our analysis can be summarized in the three broad findings described below; more specific findings and additional discussion can be found in Section 4.

**Figure 1:**
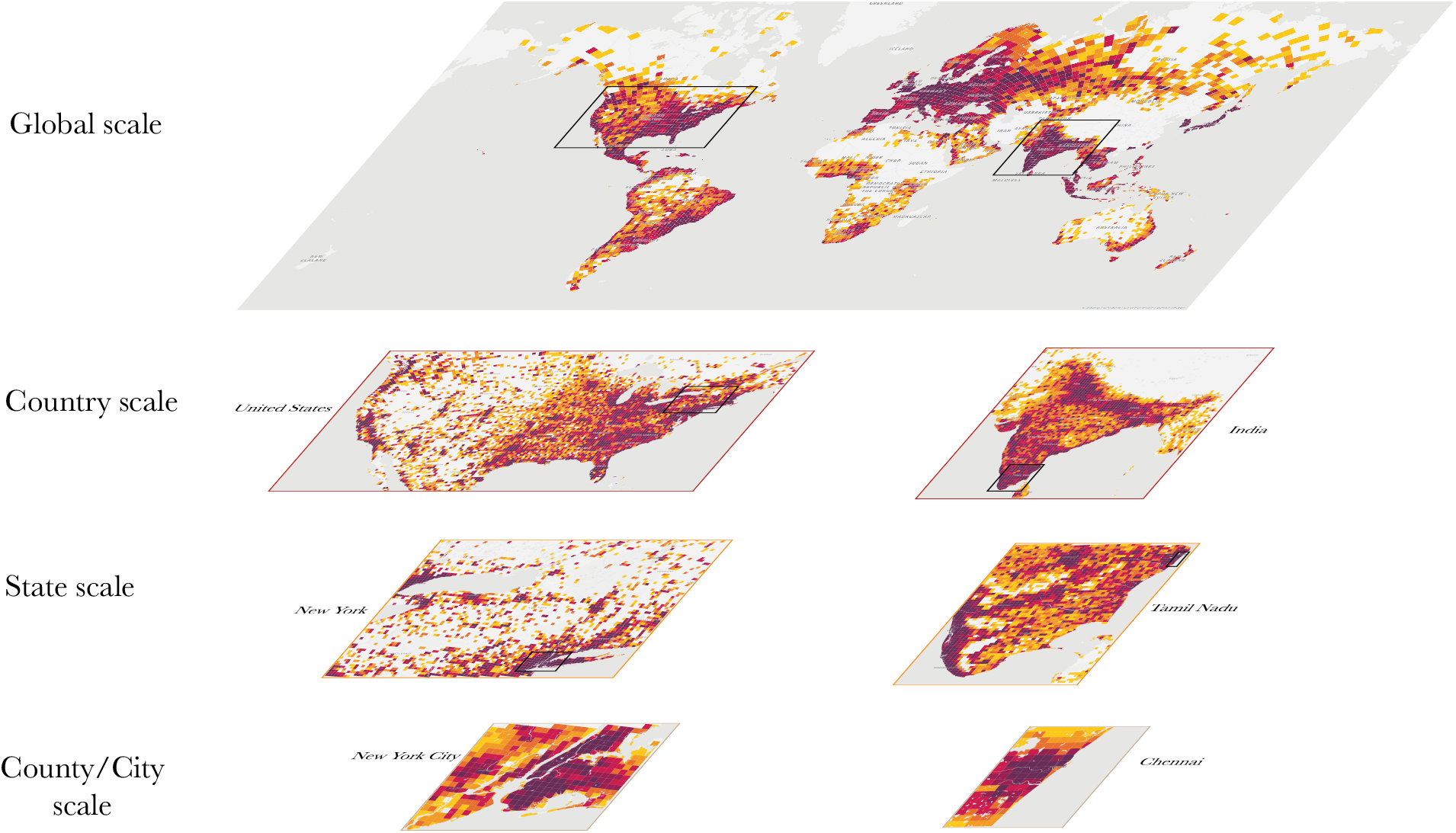
Multiple scales of mobility: mobility flows between 5 km^2^ grid cells are aggregated to appropriate spatial resolution for the analyses in this paper. The figure shows progressively the mobility volume at various S2 cell levels (L12 to L6) and geographical scales (county, state, country and globe).

In general, human mobility underwent an abrupt change in response to the COVID-19 pandemic. The mobility flows and social distancing guidelines at national, state, and county level reveal that the population complied to the social distancing guidelines and reduced mobility. Due to reopening, the population increased mobility in a relatively slow speed based on local reopening guidelines. Despite the increase in the new confirmed cases, the case growth rate decreased due to the implementation of social distancing which is partially revealed by mobility flows.

First, the global analysis reveals that the abrupt change in human mobility resulted in a 90% daily reduction in international travel between selected countries^5^, about 40-50% daily reduction of aggregate human mobility in the United States (US) and about 60-70% daily reduction of mobility in India during a 30-45 day period starting in late March and ending in late April. Since the first week of May there has been a steady increase in mobility globally. The data provides the first empirical global-scale evidence of mobility reduction and confirms various reports suggesting such a noticeable change. We observed that countries such as Japan, South Korea, and United Kingdom cut-off nearly all of its international travel in April while countries such as United States, Brazil reduced international flows by three-quarters of the baseline. In June, South Korea and Japan maintained the similar level reductions of international travels, while United States, Brazil, and United Kingdom have seen some increase in the travels, which led to 57.6%, 59.1%, 87.3% flow reductions. We also analyzed the intra-country mobility flows aggregated at country level, which can serve as a proxy for the human interactions. Compared to their January baselines, South Korea showed only 7% reduction in April, while Japan showed 30.1%. Also for United States, Brazil and United Kingdom, they are 40.0%, 41.2%, and 62.0%. These reflect the varying policies in different countries. In June, we observed alleviation of the reductions, which are 1.6%, 15.6%, 16.0%, 26.9%, 39.4% for South Korea, Japan, United States, Brazil, United Kingdom respectively.

Second, using the integrated map, we analyze the effects of both a social-distancing advisory and a lockdown using two countries, the US and India. We choose the two countries due to their size, population density, epidemiological context and differences in how policies are being implemented. Also, both countries started implementing social distancing measures at similar times (third and fourth week of March). Owing to the resolution of the data, our approach captures mobility patterns at a sub-state level (county or administrative level 2) as well. In the US, we observe a high degree of correlation between rising case counts and drop in mobility, with considerable flow reduction occurring before SAHO. In order to quantify mixing within the population, we create a metric called the social distancing index which indicates that across counties of the US, population mixing has reduced with the progression of the pandemic. We also report how inter-state mobility was significantly impacted in both US and India.

We illustrate the utility of near real-time MF by carrying out two computational experiments: (*i*) a retrospective counterfactual analysis of potential implication of a delayed lockdown and (*ii*) a prospective analysis on the effects on university reopening for in-person classes for the fall semester of 2020. In our first study, we assess the impact of delaying the lockdowns in the US and India. We find that even a one week delay would have caused substantial increase in the number of infections in both the countries. The results confirm the importance of timely lockdown in both the countries. In our second study, we use near real-time MF data, census data and data from the recent outbreak on the University of North Carolina (UNC) campus to simulate the effect of opening the University of Virginia (UVA) campus. These campuses are relatively similar. UVA has decided to delay in-class by two weeks and thus provided a good setting to study the question. The simulations indicate a potential for an outbreak at the county level.

Note that we have done our analysis based on the reported infections rather than actual infections which is unknown due to complicated testing context. The limitation of lack of accurate testing information results in bias in reported data. However, we use example to show that despite the magnitude of reported cases is smaller than actual cases, the trend of the reported cases is more likely to be consistent with the actual COVID-19 dynamic.

### 1.1 Methods

#### Data sets

**Google COVID-19 Aggregated Mobility Research Dataset**, which contains the anonymized relative MF aggregated over users within a 5 km^2^ cell. The dataset describes global human mobility patterns aggregated from over 300 million smartphone users. The data cover nearly all countries and 65% of Earth’s populated surface, including cross-border movements and international migration. This scale and coverage enable us to conduct a globally comprehensive human movement analysis in response to COVID-19 pandemic. All the flow data is aggregated to the county (administrative level 2) and state (administrative level 1) for the US and India. We refer the reader to Appendix A for a detailed description of the dataset.

**COVID-19 surveillance data** via the UVA COVID-19 surveillance dashboard [16]. It contains daily confirmed cases and death count worldwide. The data is available at the level of a county in the US and at a state level in India. Daily case counts and death counts are further aggregated to weekly counts.

#### Metrics

In order to quantify the effects of social distancing through changes in mobility, we introduce two metrics namely the Flow Reduction Rate (FRR) and Social Distancing Index (SDI). While FRR measures the reduction in *connectivity* of a region to the outside world, SDI measures the change in *mixing* within the region. Additionally we use a case growth rate (CGR) and effective reproductive number (R_eff_) to quantify the corresponding changes in case incidence, where CGR quantifies changes in observed cases and R_eff_ reflects the dynamic of expected cases that has no ground truth.

**Flow Reduction Rate (FRR)**: One way to measure the impact of social distancing is to compare the levels of connectivity before and after the SAHO/lockdowns. Given a set of nodes *V*, with flows from *i* to *j* during week *t* denoted by *f_ij_* (*t*), we first compute the average outflows during the pre-pandemic period for node *i* ∈ *V* as 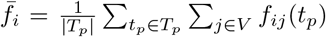 over first *T_p_* weeks of year 2020 (in our study we considered the first 6 weeks of 2020). For a given node *i*, FRR*_i_*(*t*) is then defined as

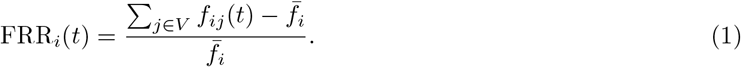

This defines a unit-less relative change in outflows from node *i* for any given week *t* with respect to 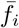. Henceforth, we omit *i* and *t* in the notation. Since `MF` is mostly symmetric (counting trips in both directions), without loss of generality we work with outgoing flows. Note that FRR is scale-agnostic and can be computed for a county, state or country.

**Social Distancing Index (SDI):** In order to quantify the *mixing* or movement within a county, we consider the flows between the 5 km^2^ cells in it. This is motivated by the fact that, under extreme case of social distancing (i.e., stay-at-home), the inter-cell flows will be significantly reduced. Let *V* denote the set of cells within a county. Let **F**(*t*) denote the normalized flow matrix of the county at week *t* with normalized flow from cell *i* to cell *j* defined as 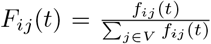. We compare **F**(*t*) to two matrices, the uniform matrix **U** and the identity matrix **I**. The uniform matrix with entries 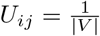, where |*V*| is the cardinality or the number of cells in *V* denotes equiprobable movement between cells and indicates a scenario of high level of mixing or movement between cells. On the other hand, the identity matrix *I*, with entries *I_ii_* = 1 and zero otherwise, indicates a scenario where all flows are within a cell and no mixing or flow happens between cells.

The SDI quantifies the closeness of **F**(*t*) to **U** and **I** and is defined as

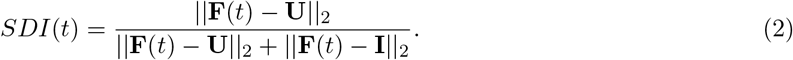

*SDI*(*t*) value close to one indicates the closeness of **F**(*t*) to the identity matrix while a value close to zeros indicates its closeness to the uniform matrix.

**Case count growth rate (CGR):** Denoting the new confirmed case count at week *t* as *n_t_*, the CGR of week *t* + 1 is computed as *log*(*n_t_*_+1_ + 1) − *log*(*n_t_* + 1), where we add 1 to smooth zero counts.

**E**ff**ective reproductive number (**R_eff_ ) [17]: This is the average number of secondary cases per infectious case in a population made up of both susceptible and non-susceptible hosts. In our study, we use an SEIR model fitted to the normalized case incidence to estimate R_eff_ as described below.

#### Impact of timely lockdown simulations

We evaluate the impact of early measures (SAHO in the US and a curfew-like lockdown in India) along similar lines as presented in [18]. As for the disease simulation, we employ a compartmental SEIR model [19, 20]. We set the disease parameters as follows: mean incubation period 5.5 days, mean infectious period 5 days, delay from onset to confirmation 11.5 days and case ascertainment rate of 15% [21]. We calibrate a weekly R_eff_ using simulation optimization to match the new confirmed cases per 100k (smoothed using Savitzky-Golay filter with filter window size of 7) at the state or country level, which is referred as the *normal scenario*. To simulate and compare the effect of a delayed intervention, we consider two counterfactual scenarios which involve *one-week delay* and a *two-week delay* in imposing said interventions. Accordingly, we extend the R_eff_ schedule by persisting a R_eff_ value in week *t* for one (or two) more weeks, and mirror the normal schedule with a one (or two) week delay. We then simulate using the modified R_eff_ schedule to produce the counterfactual simulated epicurve. Monte Carlo simulation is applied to quantify the uncertainty by adding noise to R_eff_ with noise level of AWGN with σ =0.025. We then report the number of cumulative cases avoided as of May 30 for the counterfactual scenarios. We also compare the peak time and peak values of the delay scenarios with respect to the normal scenario for United States.

#### University reopening simulations

We estimate the expected number of infected individuals coming into a university S2 cell using the MF. We first assume that the outflows from a S2 cell corresponds to the university *u*’s population *N_u_* (obtained from [22]). The fraction of flows from the counties is assumed to follow the age stratified (17-30) population distribution across US counties. Further, we divide the population based on in-state (Virginia in our case) and out-of-state population (rest-of-the-US or RoUS), that is for an university *u*,

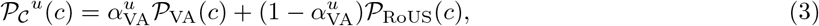

where 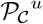 is the probability of an individual in *u* coming from *c*, 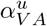 is the fraction of instate population of *u* (70% at UVA), 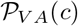 is the probability distribution of VA population and 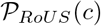 is the probability distribution of population across the rest-of-the-US counties. Given these probabilities and the *FRR*(*t*) for the cell, we can determine an estimated number of people moving to the cell corresponding to *u* as 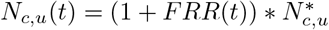, where 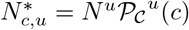.

**Expected seeding:** The possible number of infected population arriving at a cell is a function of the number of people coming and the incidence of COVID-19 in the respective counties. Hence, the expected seeding per week at a *u* is 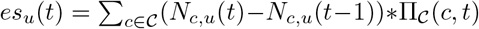, where 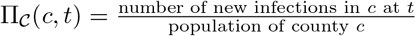 and the numerator is determined using the number of infections obtained through PatchSim simulation engine [20]. The *es_u_*(*t*) is used as the seeding of cases in our compartmental SEIR model simulation (described in the above subsection).

## 2 Results

### 2.1 Global mobility flow analysis

The evolving trend of COVID-19 dynamic differs in different localities. Thus countries’ reactions to the pandemic may present big differences. Figure 2 shows a global overview of mobility flow reduction in different countries in April and June compared to January. Considering the mobility and comparing the total outflows in January 2020 (as a baseline) with the total outflow in April, we observe that most of the countries underwent human mobility reduction, which resulted in more than 95.0% reduction for a few countries in Africa(e.g. Cameroon, Gabon, Rwanda, etc.) and a smallest 28.5% reduction in East Asia(i.e. Mongolia) in terms of inter-country mobility.

**Figure 2:**
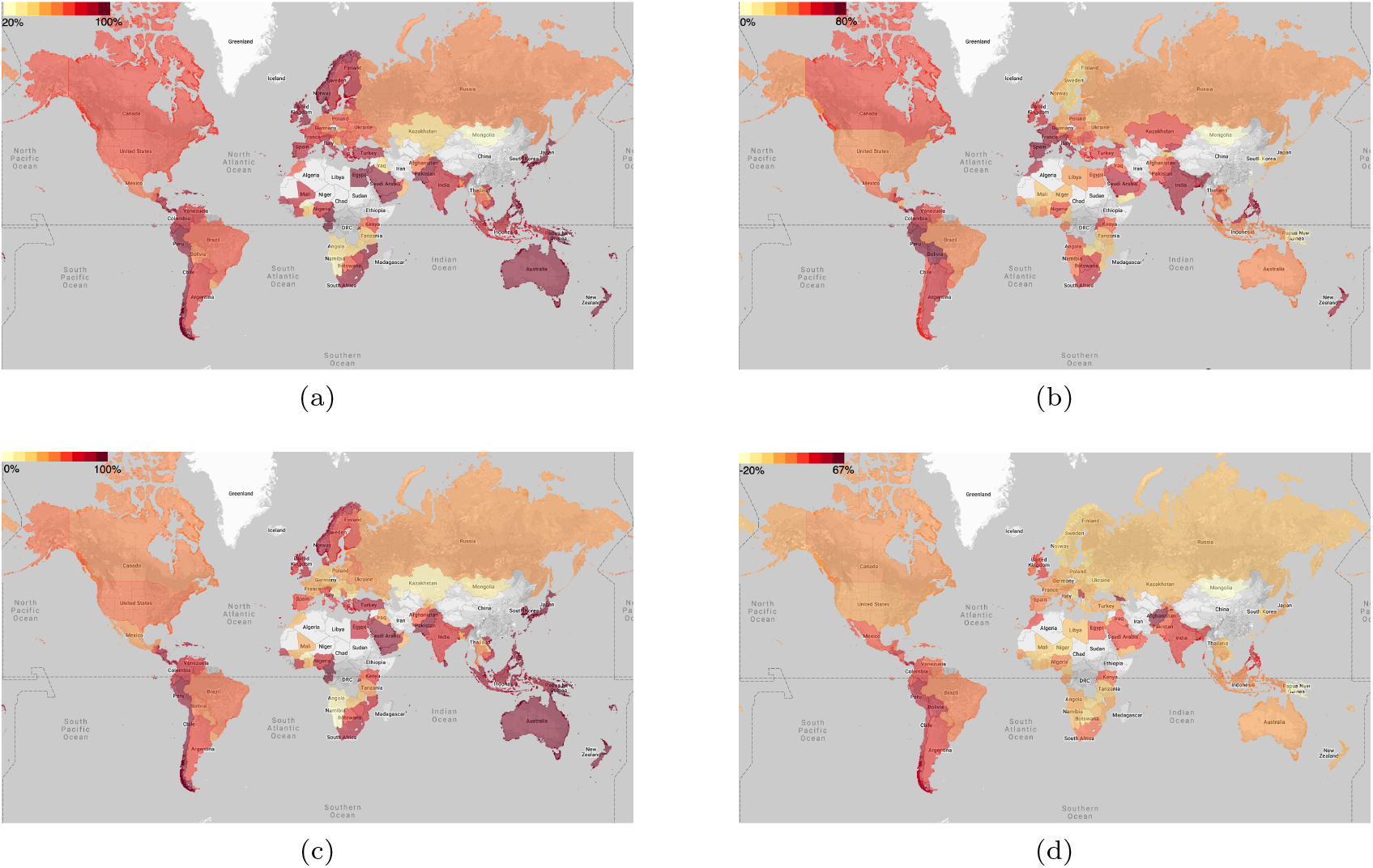
(**Global Data**) Global flow analysis: (a) Inter-country and (b) intra-country `MF` reduction across the world with flows from January and April in 2020, (c) Inter-country and (d) intra-country MF reduction from January and June in 2020. The `MF` are aggregated to country level. FRR for inter-country flow is computed with flows satisfying *i* ≠ *j*, while intra-country flow reduction FRR is computed with flows satisfying *i* = *j*. The darker the color, the stronger the flow reduction.

In order to study the impact of social distancing policies on human mobility and COVID-19 dynamics in parts of the world, we consider at least one country from each continent (except Antartica) and present their case numbers and respective MF in Figure 3. It is interesting to note that as the pandemic sets in globally, people from these countries started reducing their mobility at the same time even when there is no or few reported cases. The MF reduction in most of the countries started in the week of 2020/03/08-2020/03/14. Further, we observe that most countries show a reduction in flow a week or two *prior* to the formal lockdown announcements^6^.

Analysis of the impact of MF on the infection spread in terms of weekly new confirmed cases and cumulative growth rate (CGR) enables us to understand the interplay between mobility and cases. Towards this goal, we present MF and the weekly new confirmed case count (in Figure 3b), and MF reduction with growth of new confirmed cases (in Figure 3c). In all these countries we observe a substantial reduction in MF. The dynamics of the disease spread are not apparent from the weekly number of new cases but a consistent drop in CGR over the subsequent weeks indicates a slowing down of the spread of COVID-19 across all countries.

**Figure 3:**
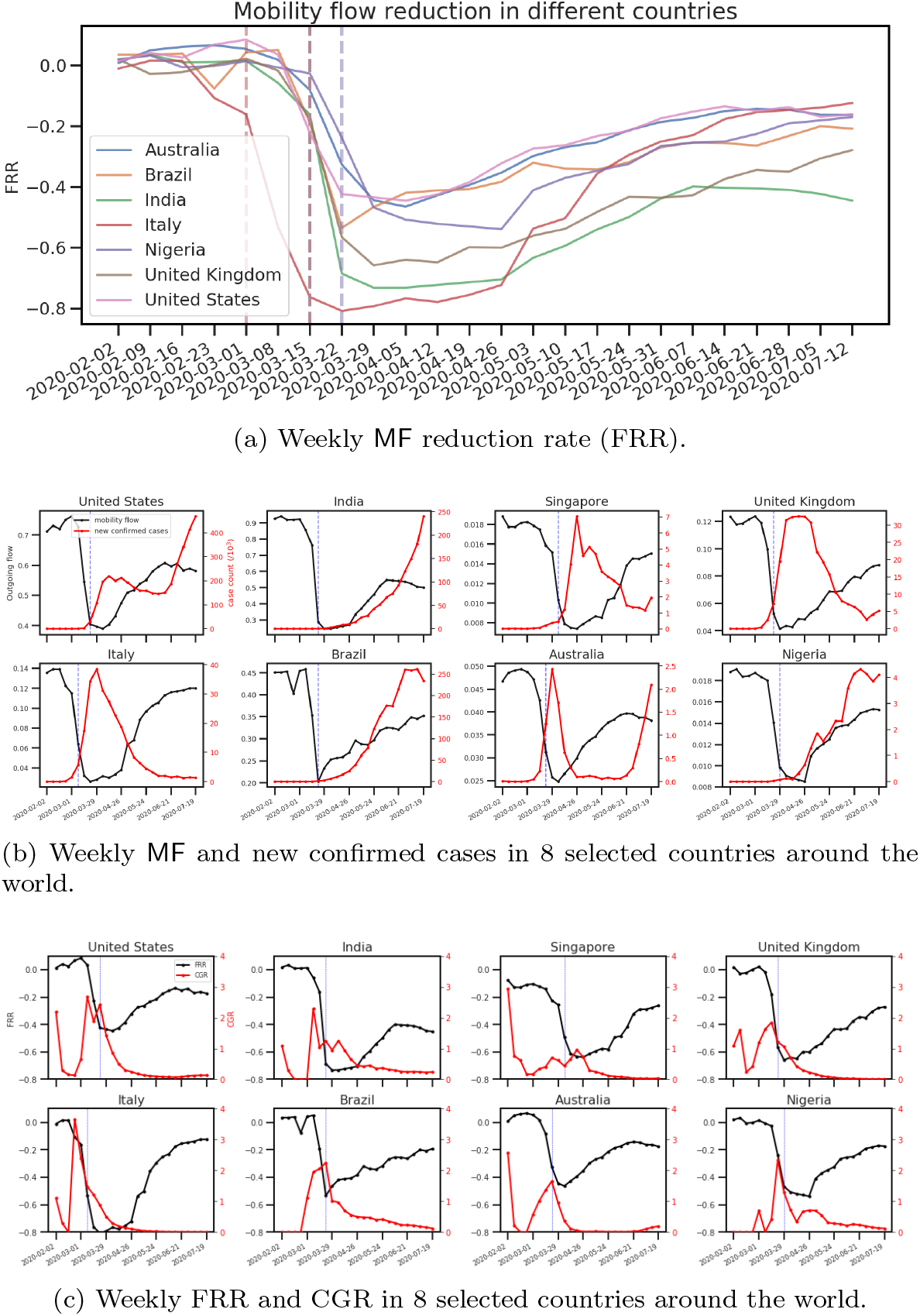
(**Global Data: A select few countries**) Impact of country wide social distancing policies on human mobility and COVID-19 dynamics. The time range is from 2020/02/02-2020/02/08 to 2020/07/122020/07/18. (a) Outgoing flow reduction from eight selected countries across the world: Australia, Brazil, India, Italy, Nigeria, Singapore, United Kingdom, and United States. To obtain these flow reduction rates (FRR), we used outgoing flows in January of each of these countries as their respective baseline and calculated the change with respect to the respective baseline from week 2020/02/02-2020/02/08 to 2020/07/122020/07/18. Each line in the plot corresponds to a country and it shows weekly FRR for that country with respect to the baseline. In addition, we use a vertical dashed line to show the week when lockdown or SAHO were imposed by each of these countries. The time of SAHO or lockdown mandates in different countries may overlap with each other. Note that FRR and lockdown lines share the same color for a given country. It is interesting to note that most countries show a decreasing trend in flow a week or two prior to the formal lockdown announcement. Moreover, most of these selected countries have initiated lockdown measures in late March 2020. (b)Weekly outgoing flows (black) vs. new confirmed cases (red) in the selected eight countries. The blue dashed lines depict the start of the lockdown week in each country. We observe that mobility flows among different countries show quite similar trends while new confirmed cases show various dynamics. But the two lines show opposite trends during February and March for all countries. (c) Weekly FRR (black) vs. CGR of the new confirmed cases (red) in the eight selected countries. The blue dashed lines depict the start of the lockdown week in each country. We observe that post-lockdown CGR is decreasing to and staying at around 0 for most of these countries despite the increase in FRR due to reopening.

We now turn to an in-depth analysis of mobility and disease dynamics for the US and India, which in many ways span the spectrum of human mobility, social distancing, government interventions, and COVID-19 dynamics.

### 2.2 Temporal analysis of MF changes with the COVID-19 progression and government social distancing orders

The temporal range is from week 2020/01/19-2020/01/25 (the first confirmed case appeared in the US) to week 2020/07/12-2020/07/18. The analysis is conducted at both state and county levels.

In order to analyze the human mobility and COVID-19 dynamics during different phases of the pandemic, we use a 4-week window and moving one week ahead each time to compute Pearson correlation between new confirmed cases and MFwithin the window. Figure 4a shows the Pearson correlation along the weeks. We observe that mobility flow and new confirmed cases show very high negative correlation (median -0.97) for almost all states during March and the correlation stays high until the mid April. The high negative correlation during March indicates that as new confirmed cases increase the mobility flows decrease. Starting from the mid April when the states started to reopen to some degree, there is a large variation in correlation values, where some are close to positive 1 while some are staying close to negative 1, which indicates that COVID-19 dynamics varies a lot due to that it is affected by multiple complicated factors like local population size, individual behaviours (e.g. wearing a mask or not in public location), and government reopening guidelines.

**Figure 4:**
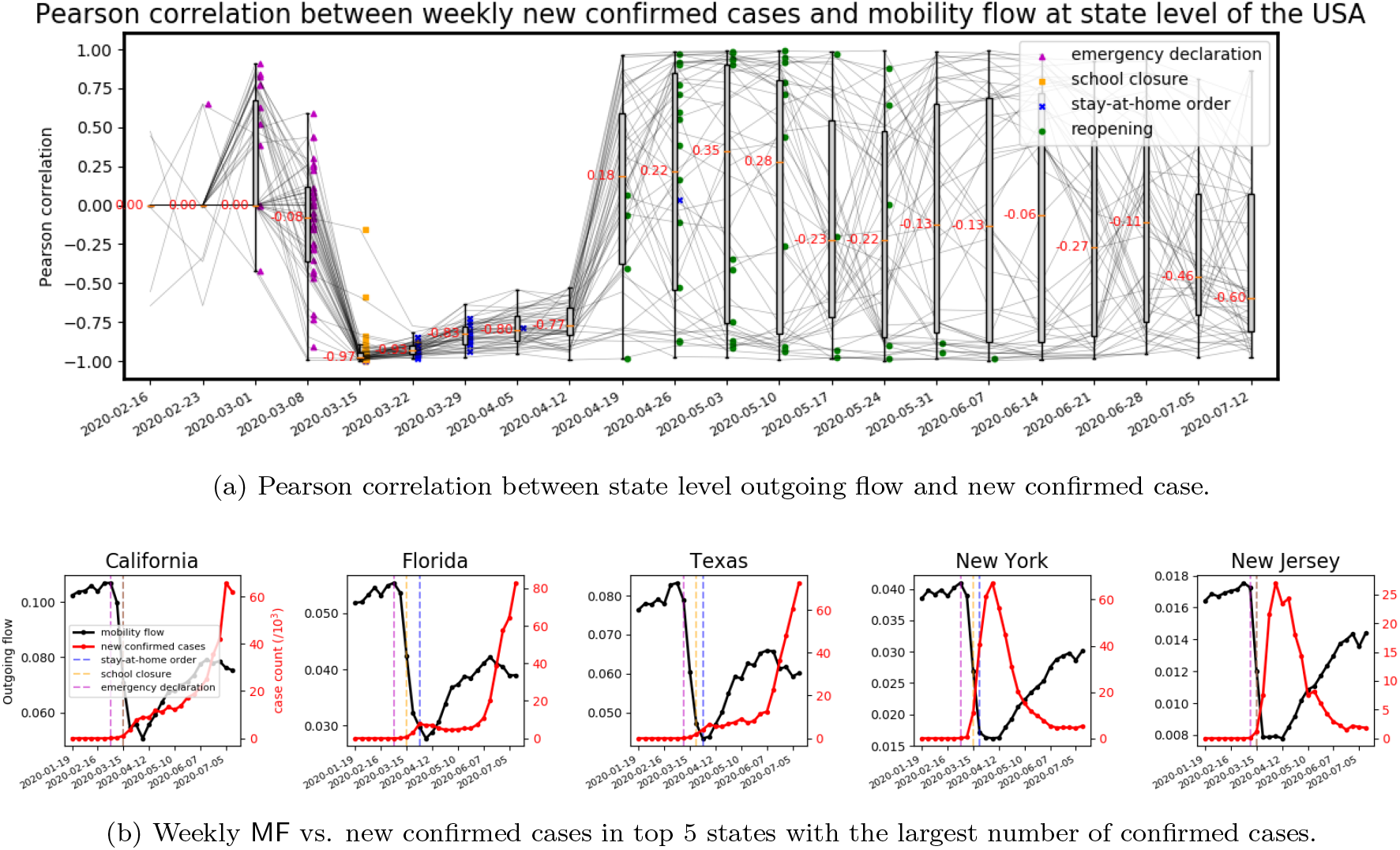
(**US Data**). Correlation analysis of human mobility and COVID-19 dynamics. (a) In order to analyze the human mobility and COVID-19 dynamics during different phases of the pandemic, we use a time series of 4-week window size to compute Pearson correlation and slide the window along the weeks. We observe that mobility flow and new confirmed cases show very high negative correlation (median -0.97) for almost all states during March and the correlation stays high until the mid of April when states start to reopen. (b) Weekly outgoing flow (black) and new confirmed cases (red) in the top 5 states with the largest number of confirmed cases. The two lines show opposite trends and cross around week 2020/03/222020/03/28. The mobility starts to drop before the SAHO (dashed blue vertical line) in 5 states.

We further explore the change in MF using the flow reduction rate (FRR) (1). Figure 5a shows a 41% (*Q*1: −50% and *Q*3: −30% in FRR) mobility reduction compared to baseline flows across the states during the week 2020/03/22-2020/03/28 where most states had declared SAHO. There is a 27% reduction in flows during the week of the most school closure order, and 18% flow reduction in the week of the most SAHO. In the subsequent three weeks after the declaration of SAH orders the flows remain nearly constant, but since then there has been a continuous rebound in the flows with flow reduction as the states reopening. The relative timing of the reduction in flows indicates that the population complied to the social distancing guidelines and reduced mobility. The largest flow reduction happened during the week of most states declared school closure orders. However the population increased mobility in a relatively slow speed according to corresponding reopening guidelines. Although social distancing orders are implemented closely in timeline by different states, the reopening process varies a lot. Figure **??** presents the timeline of the growth rate of the new confirmed cases (CGR) and social distancing orders. We observe that new confirmed cases growth rapidly in March. CGR is increasing (up to 2.11) during the early March before school closure orders while it started to drop at mid-March after school closure and SAH orders. Although states reopened gradually, we observe the CGR of all the states remain in the range of [-0.04,0.22].

**Figure 5:**
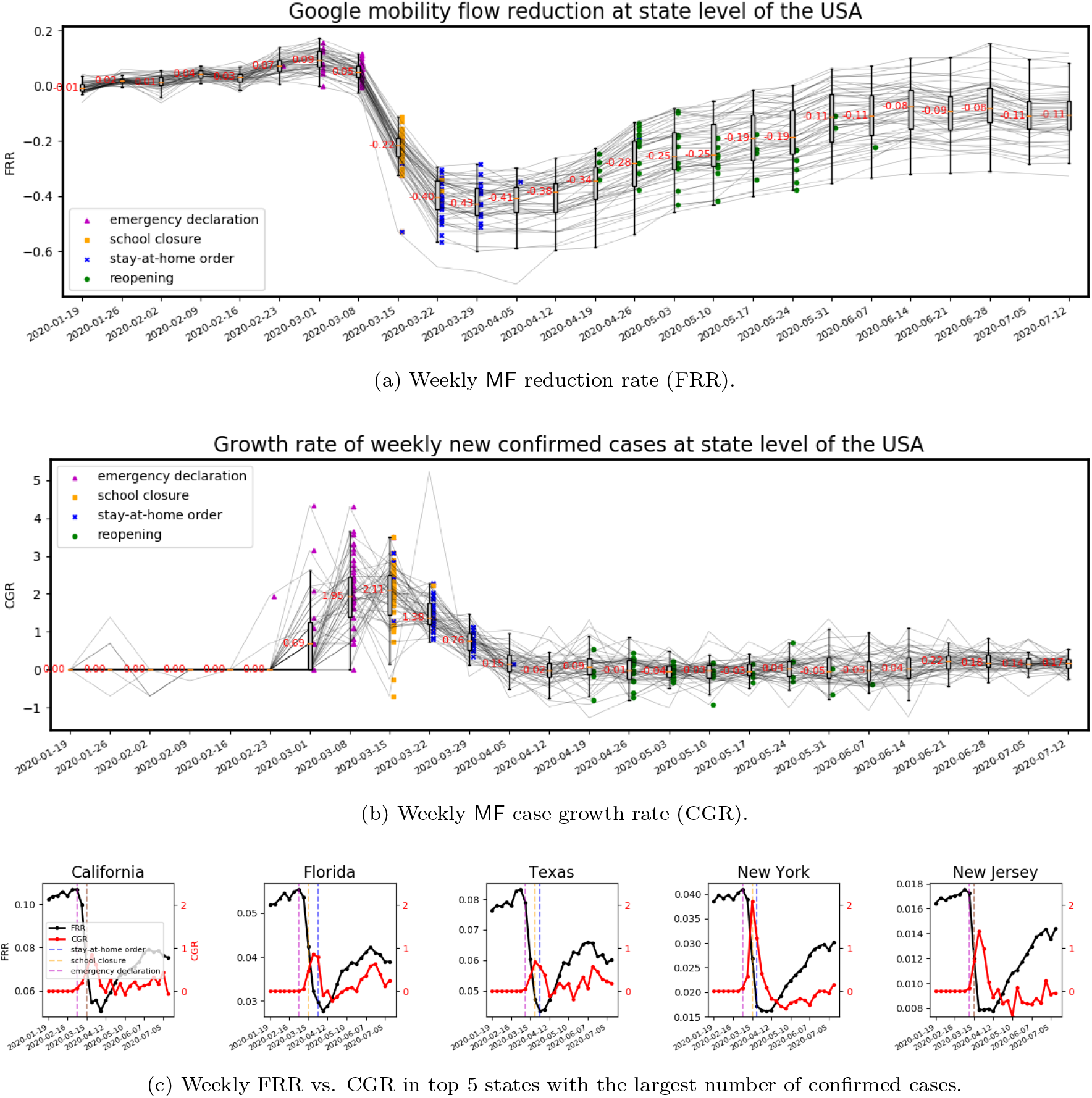
(**US Data**). Impact of state level social distancing policies on human mobility and COVID-19 dynamics. The time range is from 2020/01/19-2020/01/25 to 2020/07/12-2020/07/18. Vertical dashed lines mark the time of state level social distancing mandates including emergency declaration (purple), school closure (orange), and SAHO (blue). The time of mandates at different states may overlap with each other. (a) and (b) Spaghetti plots show the weekly FRR and CGR time series of 50 states, respectively. A boxplot is used to display variation in samples of 50 states each week. The median value is shown along with the median line (orange, also outliers are disregarded). A positive FRR indicates an increase in flow and a negative FRR means reduction in flows when compared with pre-pandemic flows. A positive CGR means the number of new confirmed cases increases and vice versa. (c) Weekly FRR (black) vs. CGR of the new confirmed cases (red) in 5 states with the highest number of confirmed cases. Although, in Figure 4b, we see that the number of new cases are increasing, (b) and (c) indicates that the growth rate of the new cases drops considerably across states.

Figure 5c presents the timeline of FRR, the growth rate of the new confirmed cases (CGR) and social distancing orders in the five states with the highest number of confirmed cases as of July 18, 2020. Although, we observe that the number of new confirmed cases increase despite the MF dropping rapidly during the week from 2020/03/01-2020/03/07 to week 2020/03/29-2020/04/04, we observe that the rate of growth of cases drops significantly. Except Florida, the CGR remains at a small value with moderate oscillation even with FRR increasing. This may due to the behaviour changes such as people started to wearing a mask.

### 2.3 Spatial distribution of mobility patterns and COVID-19 cases

Figure 6 provides a spatio-temporal view of how the FRR coincides with the state-level new cases. During the week of 2020/01/26-2020/02/01, when the US recorded its first few cases, the mobility patterns across all the states show normal behaviour. As of 2020/03/02, 100 confirmed cases were recorded across the US; by March 2020/03/17, all the 50 states had the incidence of COVID-19 and during the week of 2020/03/15-2020/03/21, New York state had recorded nearly 10,000 new cases. We observe that with the progression of the pandemic, predominantly, the most populous states have the highest number of new cases. During the week of 2020/03/15-2020/03/21 a national emergency was declared and in addition several states had already closed schools and we begin to observe an overall reduction in the mobility. We also observe that generally the states with high number of cases also have higher reduction in flow. This becomes more evident in the week of 2020/03/29-2020/04/04 where we observe flows reducing with the case counts. However, despite the continuous growth in new confirmed cases, the flow reduction started to weaken in all states in the week of 2020/05/24-2020/05/30 when most states started reopening process. The mobility kept recovering even with a surge in the number of daily new cases during the week of 2020/06/28-2020/07/04. In the interstate mobility matrix, the self-loop flows are suppressed. In order to achieve a sense of adjacency, we group the states according to their respective HHS region designations. During normal times, the inter-state mobility matrix shows considerable flows across all the states. During the week of 2020/03/15-2020/03/21 a few interstate connections start to drop and in the week 2020/03/29-2020/04/04 we observe nearly 70% of the state-pairs that are present during normal times, dropping. However, as of some of the weeks of July and August, we observe the mobility increasing substantially despite the cases increasing across states. A number of Mountain states have relatively low incidence and have shown relatively high increase in mobility.

**Figure 6:**
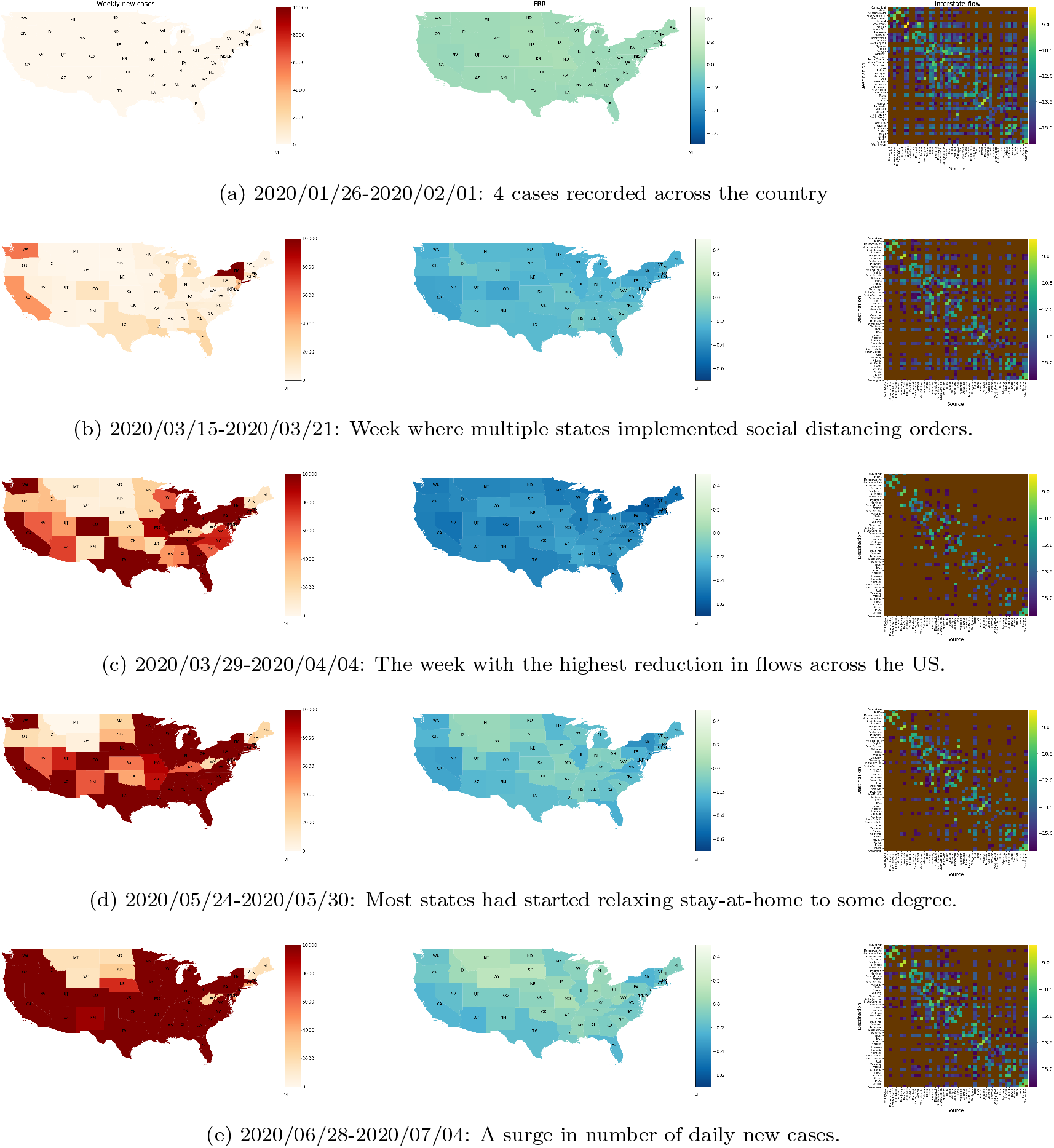
(**US Data**) A comparison of variation in number of new cases to variation in flow reduction and inter-state mobility across different weeks. Column 1 represents the spatial distribution of new cases across the US, column 2 shows the spatial distribution of FRR, and column 3 depicts the heatmap of natural logarithm of the flow between states (states are ordered as per the health and human services grouping to obtain a sense of adjacency, self-loops are suppressed and brown indicates state-pairs not recorded in the data). Row 1 represents baseline times (only a few cases recorded). Row 2 represents the week when New York and California recorded considerable number of weekly new cases and we observe that there is considerable reduction in flows across the US and also interstate mobility. Row 3 shows the week with the highest FRR across the US. By the week of 2020/03/29-2020/04/04, the weekly number of new cases has increased substantially and almost all states have brought down mobility significantly, with the number of state pairs with non-zero flows dropping by 70%. Row 4 shows the week when most states reopened to some degree. Row 5 presents the week when there is a surge in the number of daily new confirmed cases. We observe a substatial increase in mobility despite the cases increasing across the nation with the interstate mobility increasing substantially.

**Figure 6:**
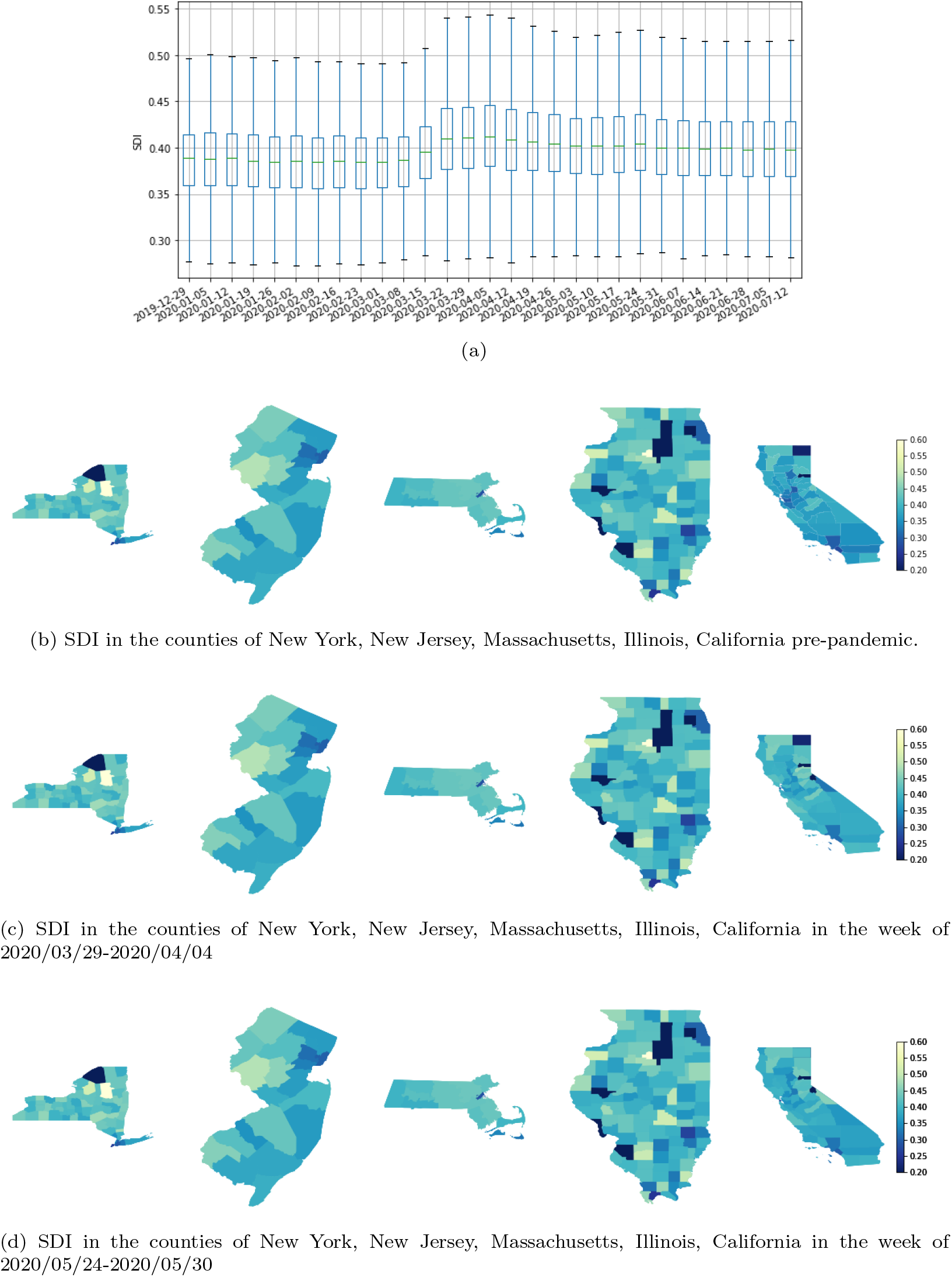
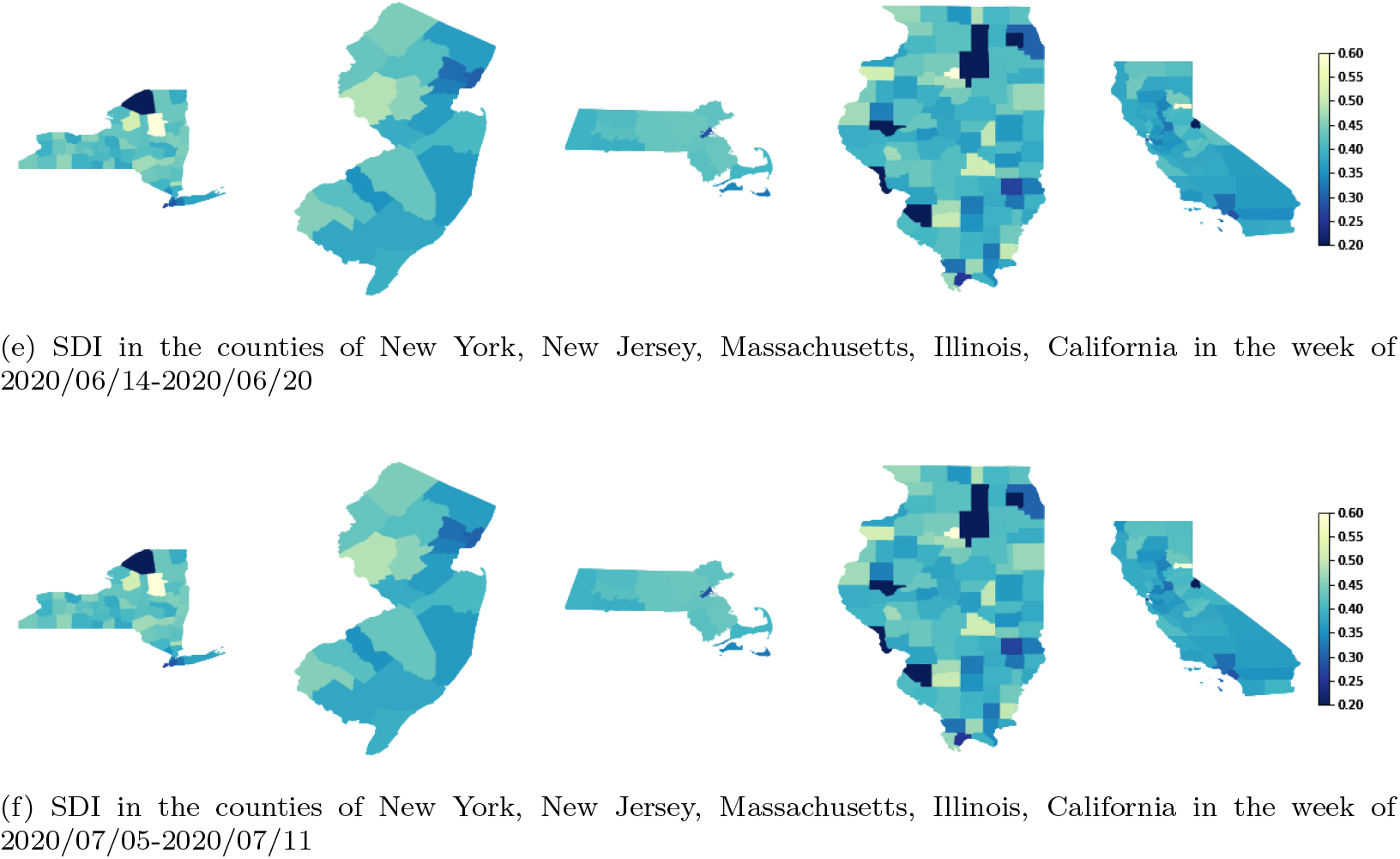
SDI at a county level. (a) Variation in SDI across all the counties of the US (obtained from the cell-to-cell flows within a county) for various weeks in the 2020. We observe that from week of 2020/03/15 (when most orders where implemented) the overall SDI starts to increase, implying that the mixing reduced. But since the last week of April we observe the overall SDI dropping and staying nearly constant over the weeks of May, June and early July. (b), (c), (d), (e), and (f) shows the choropleth plots with counties shaded according to their SDI values for five states with highest number of confirmed cases pre-and post-stateorders. Plots (d), (e), and (f) correspond to the time lines when states started phased reopening. Although, the mixing has increased, the SDI values are higher than the baseline values and across counties we do not observe much change in mixing patterns despite relaxation in social distancing orders.

### 2.4 Quantifying mixing within counties

In this analysis, we attempt to capture the mixing within a county by employing the inter-cell flows. Ideally, with social distancing orders in place, one would expect inter-cell flows within a county to drop. Due to reduced inter-cell flows, normalized flows should tend towards an identity matrix and hence *SDI*(*t*) defined in Equation (2) should move closer to 1. In Figure 7a, the boxplot represents the variation in *SDI*(*t*) across the various weeks of 2020. We observe *SDI*(*t*) to be nearly constant until the implementation of national emergency and state-level orders after which we start to observe an increase in SDI (10%). However, by the third week of April we start to observe SDI dropping and staying nearly constant over the weeks of May, which could be attributed to social distancing fatigue, a desire to return to daily routines, and other factors. Although, the SDI has reduced, the values are higher than the baseline values indicating a lower than normal mixing. As a general observation of variations in SDI across counties, we consider five states which have experienced the highest number of cases. The variations in *SDI*(*t*) across different weeks of the pandemic can be observed through the choropleth plots in Figure 7b, 7c, and 7d. We observe the overall shading moving towards yellow indicating reduction in mixing within counties. Since the last week of April, we observe a decrease in SDI indicating some degree of *social distancing fatigue* and possibly movement due to essential services. Importantly, the SDI has remained nearly constant over the month of May and median SDI nearly 5% higher than the median baseline values. A similar analysis of mobility and the disease dynamics are presented in Appendix C for India. We specifically consider India as its short range and long range mobility patterns are significantly different from the US. In addition, the population density of India is significantly higher than the US. Since the surveillance systems are evolving and the testing rates have been on the lower side, an US style analysis presented above cannot be done for India.

**Figure 7:**
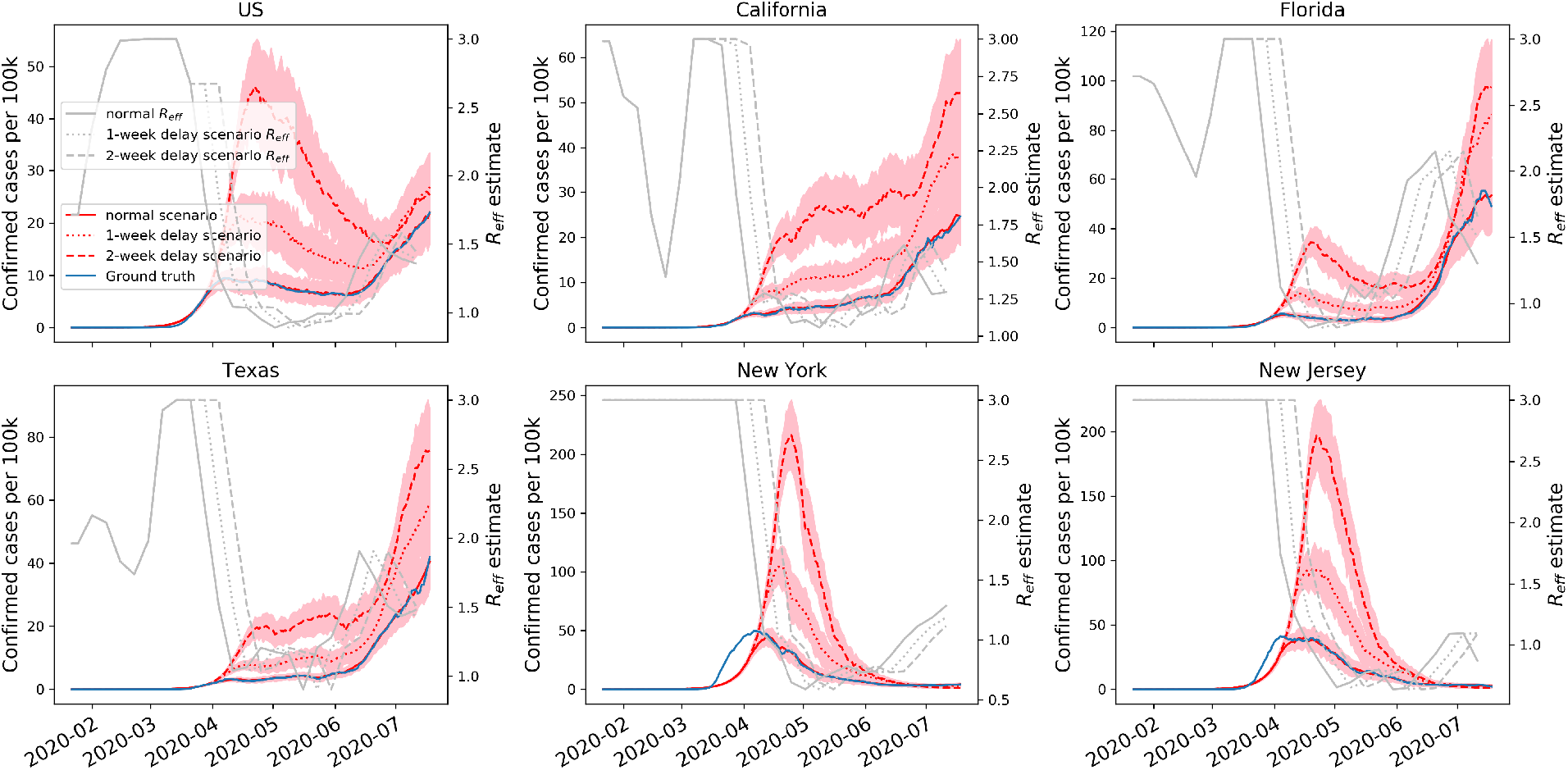
Posterior fitting to daily new confirmed cases with delayed intervention scenarios in the US and top five states with the highest number of confirmed cases. The blue line is the smoothed number of new confirmed cases per 100k. The red lines and pink shadow are the mean estimate and 95% CIs. The grey lines are fitted R_eff_ estimates. Solid lines denote normal scenario, dotted lines and dashed lines denote the scenario of one-week and two-week delay.

### 2.5 Evaluating the epidemic impact of timely lockdown

There has been considerable debate regarding the implementation strategies and timing of social distancing measures. Here we study the effects of a delayed implementation of mandates and various strategies through a compartmental SEIR model. We simulate the disease growth for two countries, the US and India. In the US, the level and timing of implementation of SAHO varied across states while in India, a curfew-like lockdown was imposed across all the states simultaneously, despite the cases localized only to a few states. Figure 7 and Figure 8, we show the simulation results. First, the R_eff_ parameter in the model is determined by fitting the smoothed daily new confirmed cases. Next, we create delay scenarios to simulate for the delayed implementation for multiple states in the US and India, where the blue line is the smoothed number of new confirmed cases per 100k. The red lines and pink shadows are the mean estimate and 95% CIs. The grey lines are fitted R_eff_ estimates. Solid lines denote normal schedule, dotted lines and dashed lines denote the scenarios of one-week and two-week delay. The numerical comparison between normal scenario and delay scenarios is presented in Table 1.

**Figure 8:**
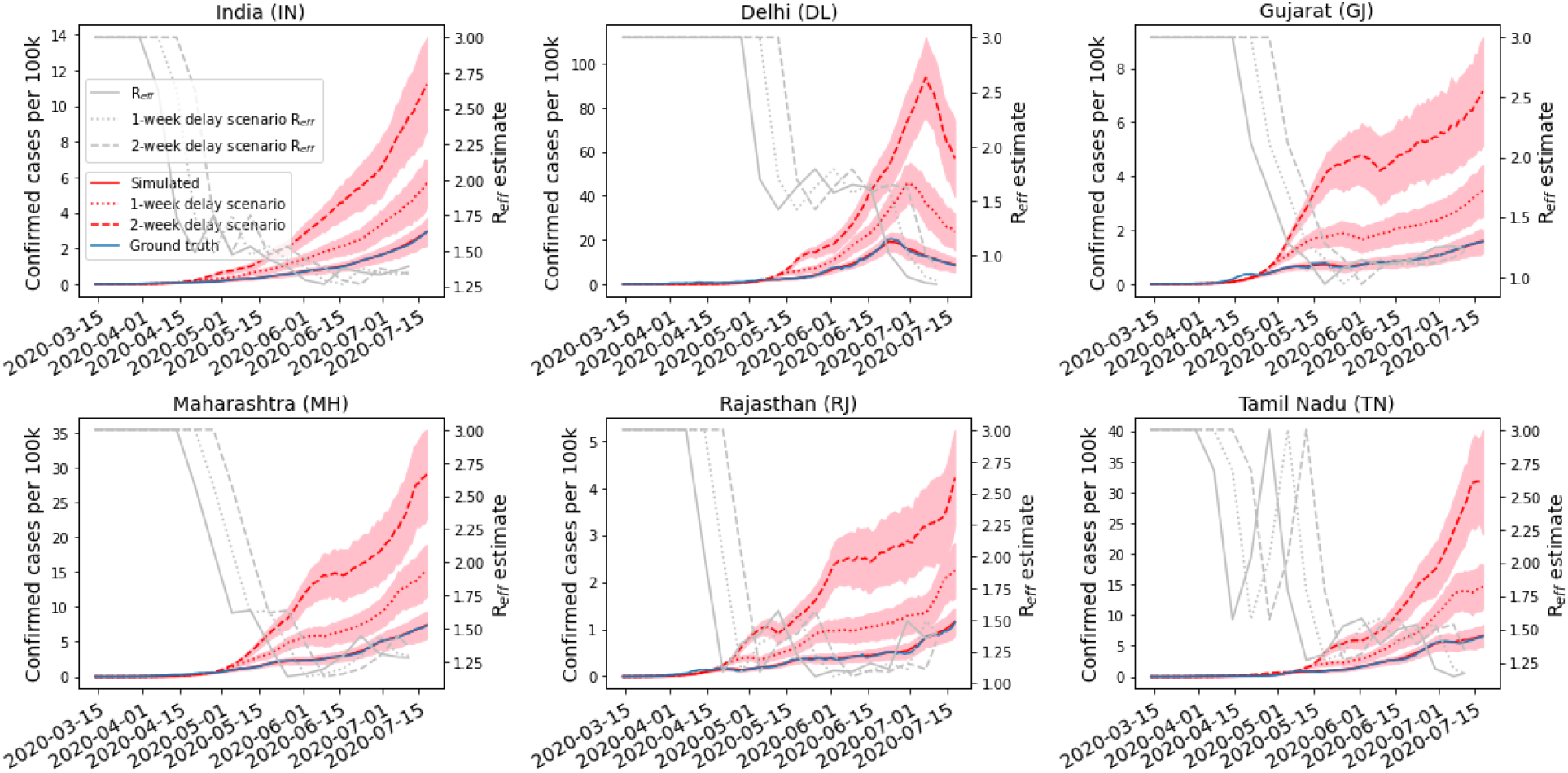
Delayed intervention scenarios for India and a few states with highest number of confirmed cases and India. The blue line indicates the smoothed number of daily cases, red lines correspond to simulation results with scenarios appropriately indicated, and the grey lines show the estimated R_eff_ and scenarios created by prolonging the R_eff_ for one week and two weeks. In general, we observe that with a one-week delay in implementation of lockdown, the total number of cases nearly doubles, while for the two-week scenario, we observe the cases increasing four folds in India. Mostly, the number of new cases is steadily increasing across India.

**Table 1:** Comparison in peak size, peak time, cumulative cases between normal scenario and delay scenarios. A ratio is given by *υ_delay_/υ_normal_* for peak size and cumulative cases, while a difference is given by *υ_delay_* − *υ_normal_* for peak time and cumulative cases. Mean values with 95% CIs are shown in the table. The peak analysis for states where peak does not appear until July 18th is done with observed maximum values.

| US |  |  |  |  |  |  |
| --- | --- | --- | --- | --- | --- | --- |
| one-week delay | US | CA | FL | TX | NY | NJ |
| peak size (ratio) | 1.25<br>[0.87,1.64] | 1.61<br>[1.09,2.12] | 1.59<br>[1.16,2.01] | 1.48<br>[1.00,1.96] | 2.37<br>[1.96,2.77] | 2.36<br>[1.89,2.84] |
| peak time (diff. in days) | -3<br>[-28,23] | -1<br>[-8,5] | 1<br>[-6,7] | -1<br>[-5,3] | 6<br>[1,10] | 5<br>[-5,16] |
| cumulative cases (ratio) | 1.69<br>[1.29,2.09] | 1.80<br>[1.39,2.21] | 1.59<br>[1.19,1.99] | 1.70<br>[1.28,2.12] | 2.05<br>[1.72,2.37] | 2.06<br>[1.74,2.38] |
| diff. in cumulative cases (K) | 2527<br>[1413,3641] | 305<br>[188,423] | 196<br>[93,298] | 224<br>[108,340] | 340<br>[261,418] | 168<br>[135,202] |
| two-week delay | US | CA | FL | TX | NY | NJ |
| peak size (ratio) | 2.22<br>[1.62,2.82] | 2.16<br>[1.51,2.81] | 1.80<br>[1.30,2.29] | 1.95<br>[1.46,2.44] | 4.79<br>[3.98,5.60] | 4.79<br>[3.94,5.65] |
| peak time (diff. in days) | -83<br>[-92,-73] | -1<br>[-7,5] | 0<br>[-5,6] | -2<br>[-6,3] | 11<br>[6,16] | 7<br>[-1,15] |
| cumulative cases (ratio) | 2.70<br>[2.12,3.27] | 3.02<br>[2.32,3.73] | 2.22<br>[1.67,2.78] | 2.66<br>[2.23,3.10] | 3.59<br>[3.10,4.09] | 3.49<br>[2.96,4.02] |
| diff. in cumulative cases (K) | 6264<br>[5019,7509] | 776<br>[597,955] | 407<br>[295,519] | 540<br>[441,639] | 846<br>[755,936] | 396<br>[353,439] |
| India |  |  |  |  |  |  |
| one-week delay | IN | DL | GJ | MH | RJ | TN |
| cumulative cases (ratio) | 1.96<br>[1.28,2.66] | 2.82<br>[1.15,4.50] | 2.23<br>[1.35,3.11] | 2.10<br>[1.57,3.17] | 1.97<br>[1.32,2.87] | 2.25<br>[1.40,3.10] |
| diff. in cumulative cases (k) | 1030<br>[785,1276] | 138<br>[109,167] | 54<br>[41,68] | 300<br>[236,365] | 30<br>[22,38] | 168<br>[132,205] |
| two-week delay | IN | DL | GJ | MH | RJ | TN |
| cumulative cases (ratio) | 3.92<br>[2.71,5.15] | 6.76<br>[2.14,3.53] | 4.6<br>[4.58,4.85] | 4.01<br>[4.53734 10.316] | 3.71<br>[3.50,7.11] | 4.89<br>[3.12,5.81] |
| diff. in cumulative cases (K) | 3147<br>[2462,3831] | 369<br>[303,436] | 174<br>[133,215] | 877<br>[691,1063] | 96<br>[74,119] | 477<br>[376 ,579] |

Figure 7 shows delay scenarios for the US and five states in the US. The search range of R_eff_ is set to [0.5,3.2] for New York and New Jersey [18, 23] and [0.5,3] for the US, California, Florida, and Texas [23], the upper bound partially to counter the effects of testing ramp-up. We postpone R_eff_ at different weeks for different states according to their SAHO. For the US, we use the week 2020/03/15-2020/03/21 when the first state level SAHO is claimed. The simulations indicate that a delay in implementation of interventions by one week in the US would have led to 2.5M [95% CI: 1.4M -3.6M] additional cases as of July 18th, 2020. If the interventions had been postponed by two weeks, the number of additional cases would have been 6.3M [95% CI: 5M -7.5M]. The ratios indicate nearly 1.69 times the ground truth number of cases for the one week delay scenario and nearly 2.7 times the ground truth number of cases for the two week delay scenario. (numbers are tabulated in Table 1).

In the context of India, the simulation results show similar effects as the US scenario. We set the searching range of R_eff_ to be [0.5,3.0] [24, 25] for the counterfactual simulation. The lockdown was implemented on the 2020/03/25 and hence, we use the R_eff_ of that week to create the one-week and two-week delay scenarios for India and all five states. The scenarios are demonstrated at both the national level and the state-level and the results are shown in Fig. 8. As indicated by the case count curves, we see an increasing trend and the peak is yet to be appear. Overall, the factor of increase in total number of cases at the national level as observed around end of July would be nearly 2 times and 4 times compared to actual confirmed cases for the one-week and two-week delay scenarios, respectively with states like Delhi and Tamil Nadu seeing a higher scaling factor. Also, the simulations project that a one week delay in imposition of lockdown would have lead to a 1M [95% CI: 785K -1.2M] additional cases while a two-week delay would have resulted in 3.1M [95% CI: 2.4M -3.8M] additional cases (numbers are tabulated in Table 1).

Although, the US has far more reported cases than India at the current time, it should be noted that the testing rate in US (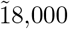 per million) is significantly higher than that in India [26].

## 3 Projecting the effects of university reopening

During early March 2020, most universities across the US called off in-person classes and resorted to online methods of instruction. With the arrival of the fall semester, many universities have been contemplating the possibility of in-person classes. We consider the University of Virginia (UVA), Charlottesville as an example to understand the effects of reopening on COVID-19 case counts in the surrounding county. UVA campus lies on the border of Charlottesville city and Albemarle county of Virginia (VA), hence, in this study, we sum the case counts and population from the two communities. A previous study investigated the effects of return of students from spring break to universities on the respective counties and compared with universities with late breaks where students returned home [27], finding a higher incidence of cases in the former case, typically two-weeks after the return of students.

We simulate the effects of reopening using an SEIR model similar to the one presented in Section 2.5. The UVA campus is relatively large (comparable to the 5km^2^ S2 cells) and hence, we consider the outflows from the S2 cells corresponding to the universities to understand the mobility patterns across the pandemic. We assume that the flows during the pre-pandemic period (month of January and February) are generated predominantly by the university population and constitute the baseline. The outflows for UVA are shown in Figure 9a where we observe a sharp decline in flow in late March 2020 after which there has been a steady increase mobility. Using the baseline population (UVA ≈ 32000) and the FRR, we compute the cell population across the weeks of 2020 as shown in Figure 9b.

**Figure 9:**
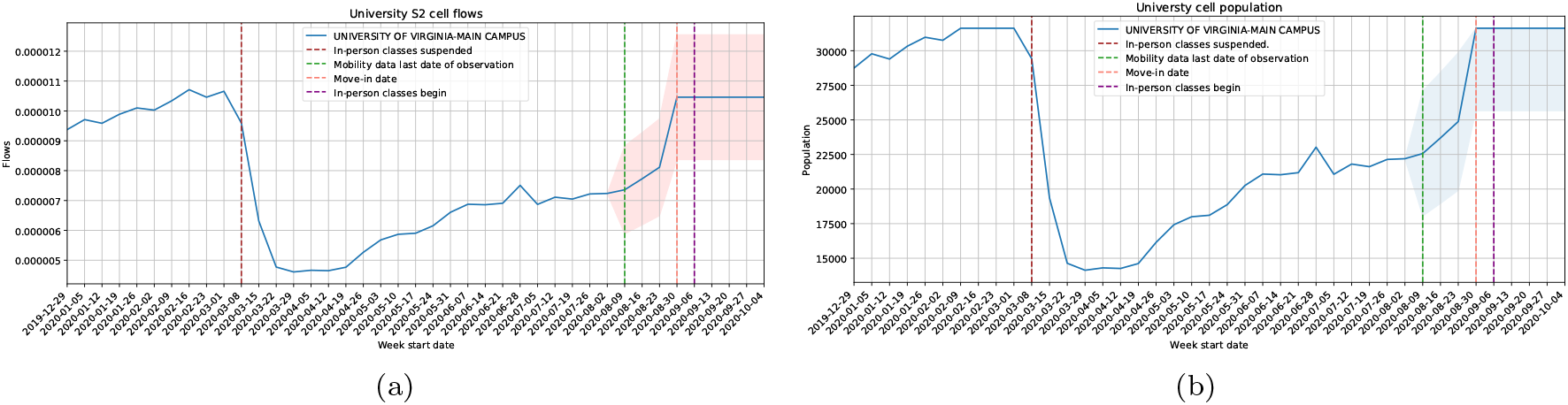
University cell flows: (a) Outflows from S2 cells corresponding to UVA. (b) An approximate amount of population in the respective S2 cells. All flows and population after the green vertical line are projections. We assume that normal flows resume from move-in date onwar.

### Flow projections and seeding

UVA has scheduled to resume classes from the September 8, 2020 and instructed students to move in starting September 3, 2020. Since the outflows are not available for future weeks, we project it forward under the assumption that there would be a steady increase of 3-7% before the week of September 3, 2020 after which all the university population would return. The hypothetical scenario corresponds to the part of the time series after the green vertical line shown in Figures 9a and 9b. With the inflow of students, we expect a fraction of them to be infected (seeds) where the fractions are determined based on county population and the incidence of COVID-19 in the respective county. The methods employed to determine the seeding of cases is detailed in Section 1.1.

### Change in R_eff_

In order to analyze the effect of reopening, we consider the recent COVID-19 outbreak experienced at University of North Carolina, Chapel Hill (UNC). UNC started in-person classes on August 10, 2020 and allowed for move-in of students on August 3, 2020. As of August 16, 2020, 130 students were reported as COVID-19 positive and UNC has suspended in-person classes since then. In order to determine the effect of the event on the county, we calibrate the SEIR model with respect to its home county (Orange County, NC) epi curve (Figure 10a). We observe a sharp increase in R_eff_ since the move-in date, which is likely in response to the reopening. We observe nearly 36% increase in the 7-day mean R_eff_ post move-in date compared week prior. Considering that the combined population of Albemarle, VA and Charlottesville, VA is very similar to Orange County, NC (≈ 150000) and the university population are comparable, we assume that the reopening of UVA would drive similar change in R_eff_. Hence, in our construction of hypothetical scenarios, we increase R_eff_ from the calibrated value by up to 30%.

**Figure 10:**
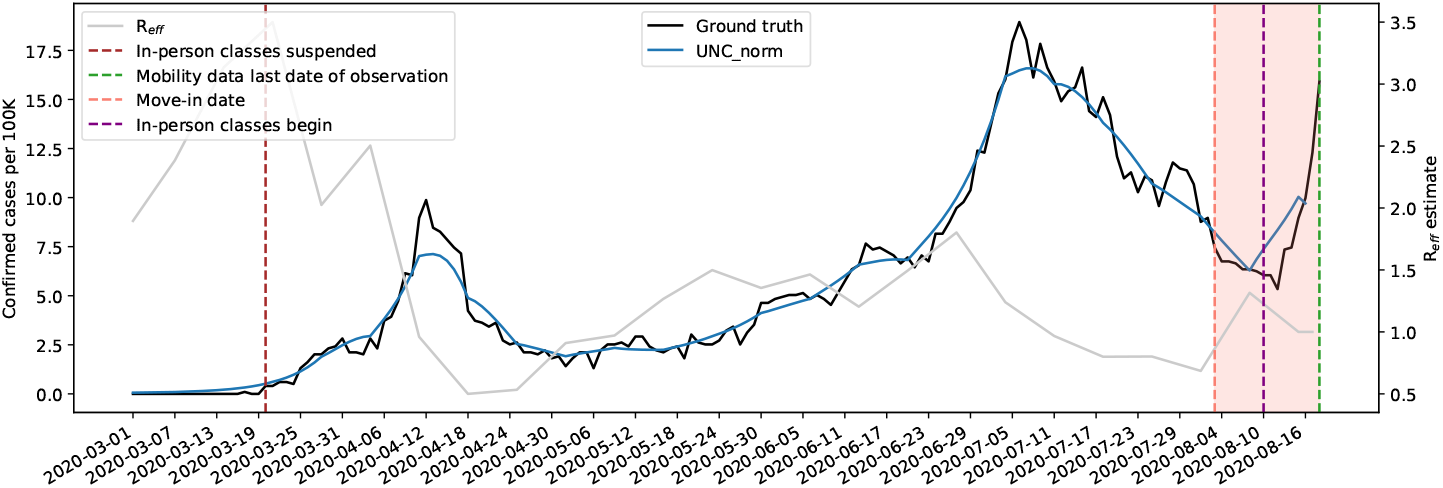
Calibration of the SEIR model over the Orange county case counts. UNC allowed move-in of students on August 3, 2020, resumed in-person classes on August 10, 2020, and reported 130 new cases by August 16, 2020. The relative change in 7-day mean R_eff_ after move-in and 7-day mean R_eff_ before move-in) was computed to be ≈30%.

### County case projections

The case projections for the county housing the university is determined using an SEIR model that is calibrated against the number of confirmed cases. The calibrated model is then run forward along with the hypothetical seeding and an experimental set of R_eff_ schedule to obtain multiple scenarios: (i) situation with reopening stalled indefinitely and hence no extra seeding where the most recent value of calibrated R_eff_ continues forever, (ii) extra seeding due to reopening and the most recent value of calibrated R_eff_ kept constant forever, (iii) extra seeding due to reopening and the most recent value of calibrated R_eff_ increases by 10% during the week of September 3, 2020 and remains constant forever, (iv) and (v) the most recent value of calibrated R_eff_ increases by 20% and 30% respectively, during the week of September 3, 2020 and remains constant forever. Scenario (ii) indicates the situation where in the classes have resumed and the social distancing norms are maintained effectively. Scenario (iii) and (iv) are used to simulate situations where in the opening has resulted in increased interaction in the county population while (v) assumes the reopening would result in similar levels of interaction as observed at UNC. We also present a situation where no seeding occurs (hypothetical situation with reopening stalled indefinitely and university population flow is minimal). The outcomes are presented in Figure 11a.

**Figure 11:**
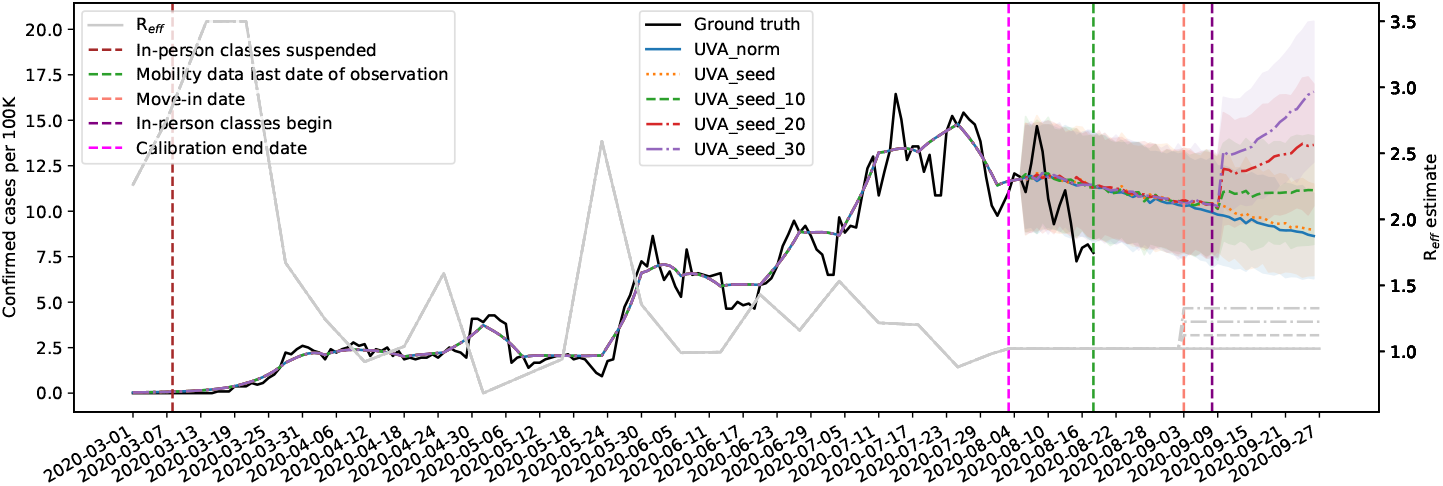
Projection of cases for Albemarle county, VA and Charlottesville county, VA combined. Scenarios ’ norm’: no extra seeding where the most recent value of calibrated R_eff_ continues forever, (ii) ’ seed’: extra seeding due to reopening and the most recent value of calibrated R_eff_ kept constant forever, (iii) ’ seed 10’, (iv) ’ seed 20’, and (v) ’ seed 30’ correspond to extra seeding due to reopening and the most recent value of calibrated R_eff_ increases by 10%, 20%, and 30%, respectively during the week of September 3, 2020 (UVA move-in) and remains constant forever. The hypothetical seeding starts from the Week of August 1, and beta is increased at the move-in date. The model is calibrated till August 4, 2020 and the projection during hold out period of two weeks suggest a relatively good fit by the model (captures the downward trend).

The results indicate that by keeping the social distancing to levels seen during the weeks prior to reopening would be effective in mitigating the effects of extra seeding of cases and one could expect 442 [95% CI: 333-551] cases over a month. This reflects the fact that Virginia has a relatively low prevalence numbers overall. The effect of seeding gets pronounced when the levels of contact increase – an increase in R_eff_ at the move-in date see a surge in cases a week later. An increase in R_eff_ by 10% would result in 469 [95% CI: 337-601] cases, an increase by 20% could lead to 498 [95% CI: 356-639], while an increase by 30% could result in 530 [95% CI: 364-697] cases over the month in Charlottesville and Albemarle counties. In order to validate the performance of the SEIR model, we calibrate the model using data up to August 3, 2020 and hold out the last two-weeks of the most recent data (green line) for testing. We observe that the projections closely track the ground truth data during the hold out period and captures the downward trend in daily new case counts per 100K.

## 4 Discussion

With the findings at global, national, state, and county level, we have consistent observations that human mobility reduce due to government social distancing guidelines. The reduction happens to long and short term travels including intra/inter-country, intra/inter-state, and intra/inter-county mobilities. Informed by world wide cases information, most of countries implemented lockdown orders in a similar timeline (start in early March) thus resulted in an abrupt mobility flow reduction at early March. By investigating state level mobility flow and COVID-19 dynamics in the US and India, we again observe relatively consistent mobility trends but with different magnitude despite that the COVID-19 evolved quite differently in two countries. Looking at details of state level interplay of MF and COVID-19 dynamics, although we observe the number of new confirmed cases increase as the mobility flow reduce, the growth rate of the new cases drops considerably across states. Even with the MF rebounding in late April due to the reopening of some states, the growth rate of the new cases remains steadily low around zero. Partly, this may be due to the better adherence to guidelines like maintaining 6 feet distance and wearing a face mask at public locations by individuals. The implication of varying levels of strictness in implementation and execution of social distancing measures was studied using US and India. The simulations project that the total number of cases with a one-week and two-week delay in interventions would have been nearly 2 times and 3 times the actual confirmed cases in the US. In India, a delay in the execution of the centrally mandated lockdown could have lead to nearly 2 and 4 times the number of cases observed. Another scenario of particular interest is the university reopening and its consequences as fall semester approaches. Using the data from the outbreak experienced at UNC, we observe a significant increase in R_eff_ of the respective county at the time of reopening. One could speculate that the increase in R_eff_ could be much higher than the increase in county R_eff_. The simulations suggest that an increase in R_eff_ and extra seeding of cases could lead to an outbreak and with the fall semester mostly synchronized across the US, the effects of reopening could be significant.

### Testing analysis

Our interplay analysis is based on the publicly available data on COVID-19 case counts that are marred by inconsistent and non-standardized testing strategies. The testing data is obtained from The COVID Tracking Project [28] and attempts to provide the Test Positivity Rate (TPR) across the US which according to WHO is a good indicator of the extent of testing. A positivity rate of 5% or below typically indicates that a state has sufficient testing capacity compared to the size of the outbreak. A Higher positivity rate typically suggests the lack of widespread testing for capturing community-level transmissions and restricted restricted to patients presenting severe symptoms. Ideally, the positivity rate should be the number of people who test positive divided by number of people who are tested, however, some states include in their testing duplicate counts (multiple testings done on an individual), as well as antibody tests (serology). Hence, the true number of positive cases is not captured in all cases. In addition, many states are unable to track the number of people tested and track only the number of tests. Hence, The COVID Tracking Project defines the positivity rate as the number of cases divided by number of negative tests plus number of cases to be consistent across states.

We use specific examples to observe the spatial (across states) as well as the temporal variations in testing positivity rates with the daily values aggregated to weekly values. *Testing rate per 100K* quantifies the number of people getting tested per 100K population. Figure 12 shows COVID-19 dynamics (new confirmed cases per 100K and CGR) and testing trends (testing per 100K and TPR). The second row presents the TPR which we observe is noisy due to that the testing capacity and testing guidelines changing at different stages.

**Figure 12:**
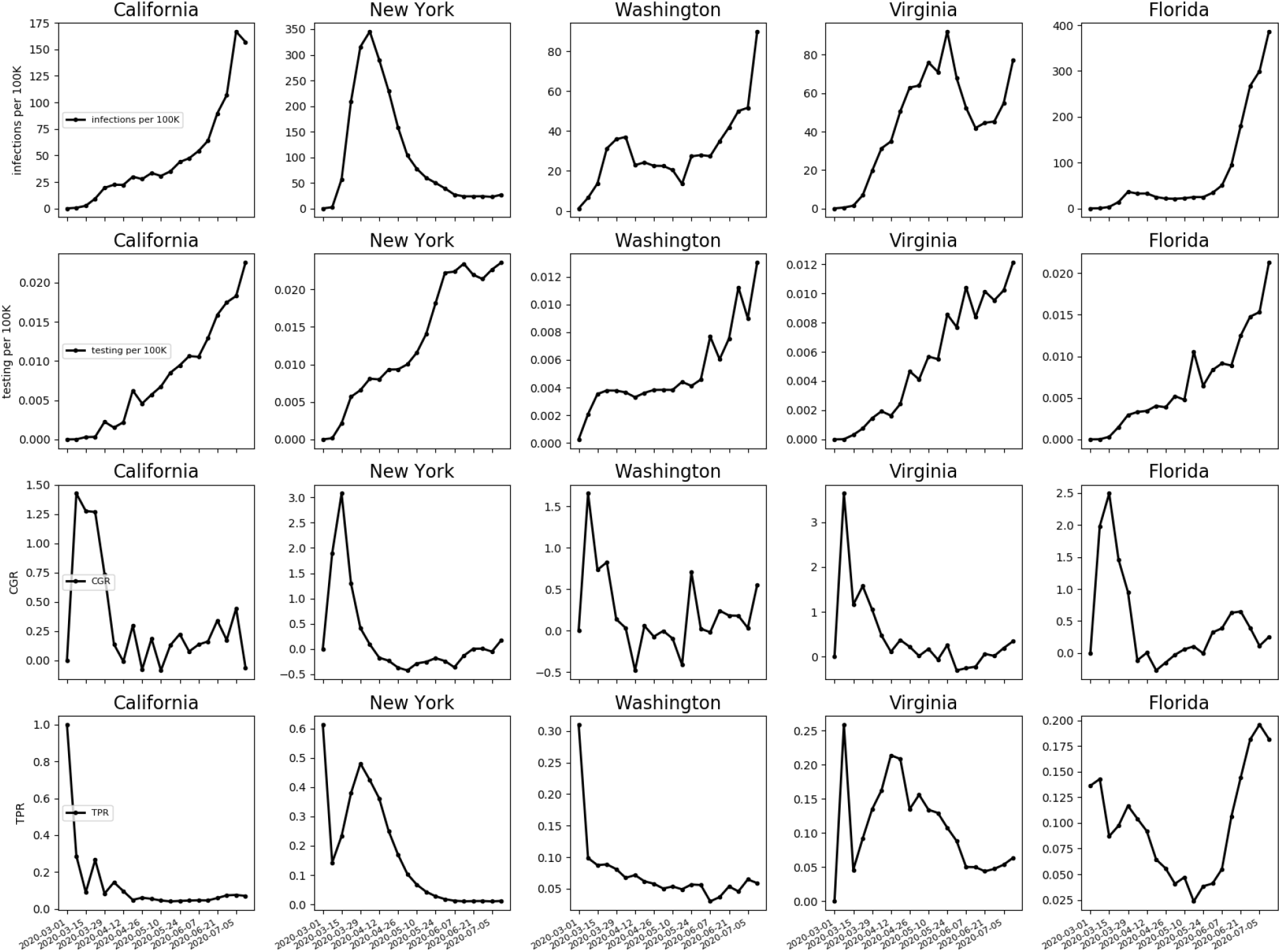
Examples of COVID-19 dynamics and testing trends of five states in the US. The first and second rows show new confirmed cases per 100K and testing per 100K. The third and fourth row show CGR and TPR.

The TPR plots (second row in Figure 12) show high values of TPR in March during which time the tests were mostly limited to individuals with COVID-19 symptoms and hence does capture the true levels of incidence in the population. In the subsequent weeks we observe that the testing rate constantly increasing thus indicating a larger coverage of population (people with mild symptoms or asymptomatic individuals).

During this phase (especially during June and July), we observe that California, New York, and Washington observe TPR stabilizing. Thus the increasing trends of new reported cases in California and Washington might be due to the increase in testing rate, while the number of new reported cases in New York has decreased dramatically even with an increase in testing rate, which indicates a true decrease trend in New York. As for Florida where the number of tests are ramping up, there is sharp increase in the TPR, indicating a true increase in the number of cases.

Given the limited information, we believe that the actual number of confirmed cases is higher than the reported values due to the testing capacity in the US and a sharp uptick in trends of reported cases could be due to the increase in the testing rate (which is further increased if there is a higher disease prevalence in a region). Hence, the reported trends, to a large degree, reveal the actual COVID-19 dynamics.

## 5 Limitations

These results should be interpreted in light of several important limitations. First, we have already discussed the limitations due to lack and non-uniformity of testing data. Second, the Google mobility data is limited to smartphone users who have opted in to Google’s Location History feature, which is off by default. These data may not be representative of the population as whole, and furthermore their representativeness may vary by location. Importantly, these limited data are only viewed through the lens of differential privacy algorithms, specifically designed to protect user anonymity and obscure fine detail. Moreover, comparisons across rather than within locations are only descriptive since these regions can differ in substantial ways. Third, the retrospective and prospective simulations are based on multiple assumptions motivated by related and relevant events; nevertheless the results provide good worst case bounds. Important assumptions include (i) relative ratios of asymptomatic to symptomatic cases is as reported in the literature, (ii) that the incidence rate in the overall population is the same as that in the student population and (iii) Universities have not put in place strict testing regimes on the campus.

### Data sharing statement

The Google COVID-19 Aggregated Mobility Research Dataset used for this study is available with permission from Google, LLC. University of Virginia COVID-19 Dashboard that provides detailed, global epidemic surveillance data and used in this study is available for public use on https://nssac.bii.virginia.edu/covid-19/dashboard/. The data on social distancing guidelines in US states and India COVID-19 surveillance data would be shared upon request and would be uploaded on https://dataverse.lib.virginia.edu/

## Data Availability

Yes, if requested, we would provide the data employed in this manuscript

## Acknowledgements

We thank Aaron Schneider, Aaron Stein, Ahmed Aktay, Alvin Raj, Amy Chung-Yu Chou, Andrew Oplinger, Ashley Zlatinov, Blaise Aguera y Arcas, Bryant Gipson, Charina Chou, Christopher Pluntke, Damien Desfontaines, Eric Tholome, Ewa Dominowska, Gregor Rothfuss, Iz Conroy, Janel Thamkul, Janet Whiteman, Jason Freidenfelds, Jeff Dean, Karen Lee Smith, Katherine Chou, Leeron Morad, Lizzie Dorfman, Marlo McGriff, Mia Vu, Michael Howell, Paul Eastham, Rif Saurous, Rishi Bal, Royce Wilson, Ruth Alcantara, Shawn O’Banion, Stephanie Cason, Thomas Roessler, Vivien Hoang, Yanning Zhang, Manish Gupta, Pankaj Gupta, Ashwani Sharma and Divy Thakkar for their support and guidance. We also thank members of the Biocomplexity Institute and Initiative, University of Virginia for useful discussion and suggestions. This work was partially supported by National Institutes of Health (NIH) Grant 1R01GM109718, NSF BIG DATA Grant IIS-1633028, NSF DIBBS Grant ACI-1443054, NSF Grant No.: OAC-1916805, NSF Expeditions in Computing Grant CCF-1918656, CCF-1917819, US Centers for Disease Control and Prevention 75D30119C05935, DTRA subcontract/ARA S-D00189-15-TO-01-UVA, and a collaborative seed grant from the UVA Global Infectious Disease Institute.

## Appendices

### A Google COVID-19 Aggregated Mobility Research Dataset

The dataset contains anonymized mobility flows aggregated over users who have turned on the Location History setting, which is off by default. This is similar to the data used to show how busy certain types of places are in Google Maps — helping identify when a local business tends to be the most crowded. The dataset aggregates flows of people from region to region, which is here further aggregated at multiple geographical resolutions weekly.

To produce this dataset, machine learning is applied to logs data to automatically segment it into semantic trips [29]. To provide strong privacy guarantees, all trips were anonymized and aggregated using a differentially private mechanism [30] to aggregate flows over time. This research is done on the resulting heavily aggregated and differential private data. No individual user data was ever manually inspected, only heavily aggregated flows of large populations were handled.

All anonymized trips are processed in aggregate to extract their origin and destination location and time. For example, if users traveled from location *a* to location *b* within time interval *t*In assessing public levels of compliance to social distancing, several analyses have revealed reduction in overall mobility.

### B Inter-and Intra-country mobility flow reductions

The magnitudes of reductions in the inter-county mobility flows become weaker for all continents, except Oceania which maintains a similar strength. As shown from 2, Europe experienced the highest drop in the averaged mobility reduction across all countries in the same continent, from 77.1% in April to 49.8% in June. For intra-county mobility flows, the averaged reductions per continent are alleviated in June for all continents, and it is most significant for Oceania with 40.1% in April to 6.1% in June.

**Table 2:** Statistics about the reductions in inter-country mobility flows comparing April and June to January, respectively.

| Continent | AVG in April | AVG in June |
| --- | --- | --- |
| Africa | 74.9% | 63.5% |
| Asia | 81.1% | 73.7% |
| Europe | 77.1% | 49.8% |
| South America | 82.0% | 72.1% |
| North America | 92.0% | 85.9% |
| Oceania | 99.6% | 99.9% |

**Table 3:**
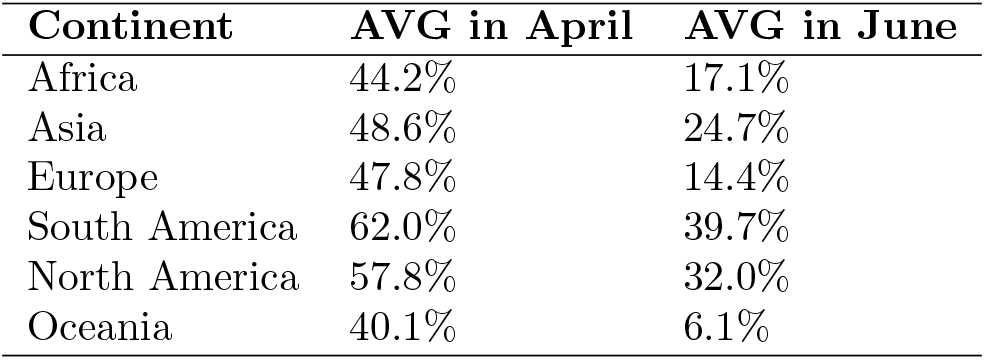
Statistics about the reductions in intra-country mobility flows comparing April and June to January, respectively.

### C Analysis of mobility flows in India and its correlation with cases

The temporal range is from week 2020/03/01-2020/03/07 (the first confirmed case appeared in India) to week 2020/07/12-2020/07/18. The analysis is mainly conducted at the state level in India due to lack of authoritative infection data at district level.

Figure 13b shows Pearson correlations between weekly outgoing flow and new confirmed case count at the state level (the date range restricted from 2020/03/01-2020/03/07 to 2020/05/03-2020/05/09 to capture the initial phase of the pandemic). We observe that the correlation varies across states and shows high (−0.75 in Uttarakhand) to low (−0.27 in Punjab) negative correlation between MF and new confirmed case count. We have ignored smaller states with less than 10 COVID-19 cases from this analysis.

**Figure 13:**
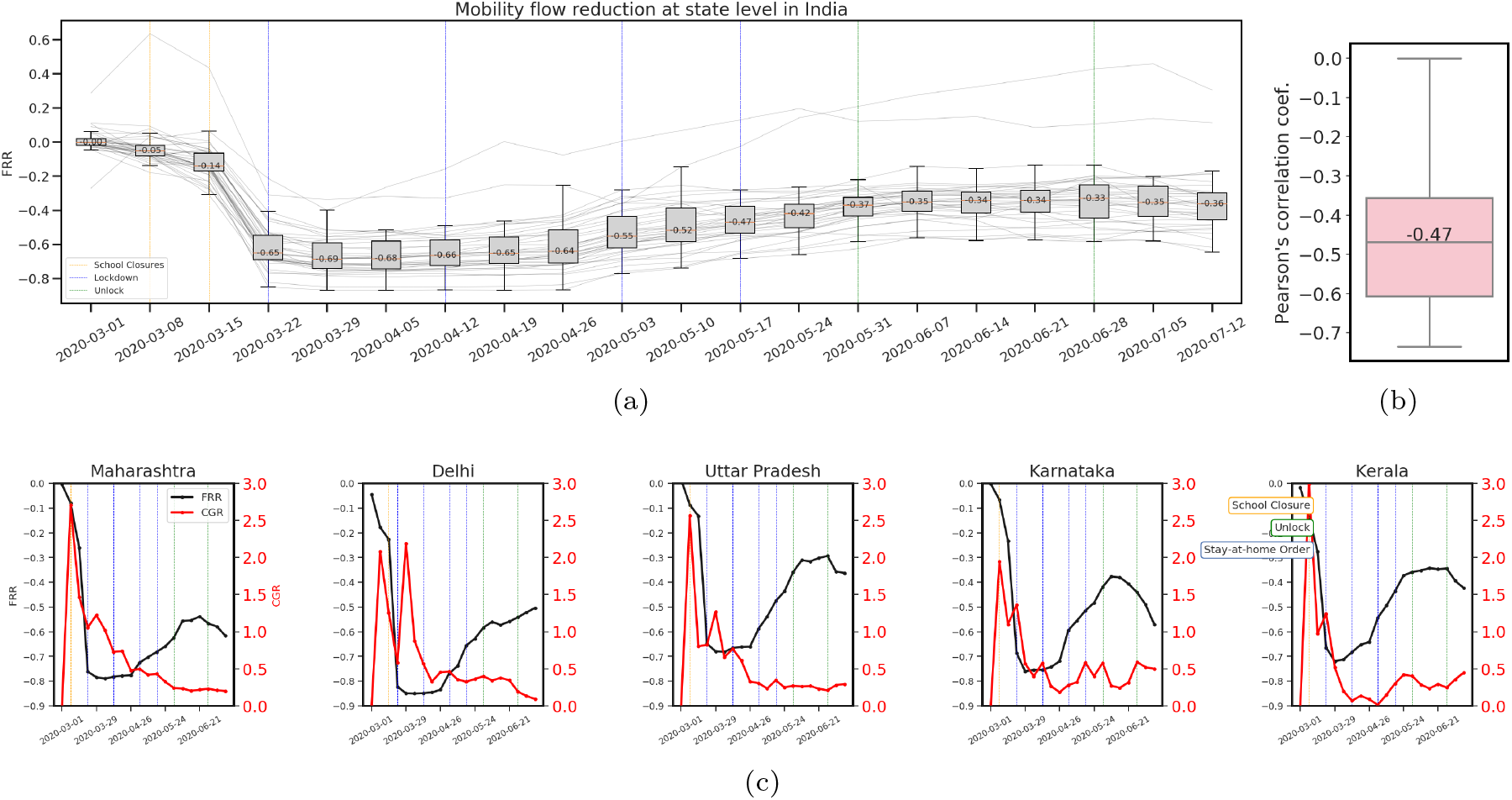
(**India Data**) Impact of state level social distancing policies on human mobility and COVID-19 dynamics. The time range is from 2020/03/01-2020/03/07 to 2020/07/12-2020/07/18. Vertical dashed lines mark the time of state level social distancing mandates including school closure (orange), lockdown (blue) and unlock (green) orders. The time of different mandates at different states may overlap with each other. (a) A spaghetti plot showing the weekly FRR time series for the 28 states and 8 union territories. A boxplot is used to display variation in samples of each of these entities every week. The median value is shown along with the median line (orange). A positive value means flow increase compared with pre-pandemic flows, while a negative value means a reduction in flows. We observed significant reduction in MF after the on-set of COVID-19 pandemic in India. The lockdown measures further reduced the flow. Towards the end of second lockdown (May 3, 2020), we observe slight increase in the MF and since then there has been a steady increase in flows. By mid-July, the MF reduction stands at 36% compared to the pre-pandemic time. (b) Distribution of Pearson’s correlation coefficient between state/union territory level outgoing flow and new confirmed case count at weekly levels during the initial phase of the pandemic. Note that we have only considered states with total cases more than 10 for this analysis. We observed Pearson’s correlation coefficient of -0.47 for India, which indicates a moderately negative correlation between the outgoing MF and newly confirmed cases per week. (c)Weekly FRR vs. CGR in top 5 states in India: the national capital state (Delhi), financial capital state (Maharashtra), IT state (Karnataka), the most populous state (Uttar Pradesh) and state with early outbreak (Kerala).

We further explore the MF change by applying flow reduction rate (FRR) (1). Figure 13a shows a 65% (*Q*1: −69% and *Q*3: −57% in FRR) mobility reduction compared to normal flows across the states during the week 2020/03/22-2020/03/28 where the central government had declared the first lockdown. Many states across India, large and small, like Maharashtra, Karnataka, Uttar Pradesh, Kerala, Uttarakhand, Odisha, Bihar, Chattisgarh, Punjab and Manipur declared school closure in the week 2020/03/08/-2020/03/14. Delhi announced school closure a week after these states. We observed 10% flow reduction in the next week after the school closure orders in the week 2020/03/08-2020/03/14. In the following week 2020/03/152020/03/21, many corporations issued work from home advisories and government offices decided to function with reduced staff strength with rotation. This seems to have resulted in a significant drop in the flow by 51%. The people’s curfew on March 22, 2020 followed by nationwide lockdown for 21 days starting March 25, 2020 further reduced the flows by 2%. These flow levels were maintained for the next couple of weeks till the end of the first lockdown. The second lockdown in India was taken for 19 days between April 15, 2020 to May 3, 2020. We observe a slight increase in flows in the first week of the second lockdown and an increase of 10% in the third week. This increase can be attributed to allowance of certain economic activities in less affected areas post April 20. Over the month of May, the flows have steadily increased by nearly 40% compared to flows during the imposition of lockdown. Thus the lockdown orders and graded resumption of economic activities seems to have large impact on mobility in India.

The effect of lockdown orders on mobility can be observed by comparing inter-state flows prior (Figure 14b), post lockdown (Figure 14c) and during unlock phases (Figure 14a, Figure 14b). The usual flow of inter-state mobility has either reduced significantly or dried up completely (e. g. Uttar Pradesh-Madhya Pradesh).

**Figure 14:**
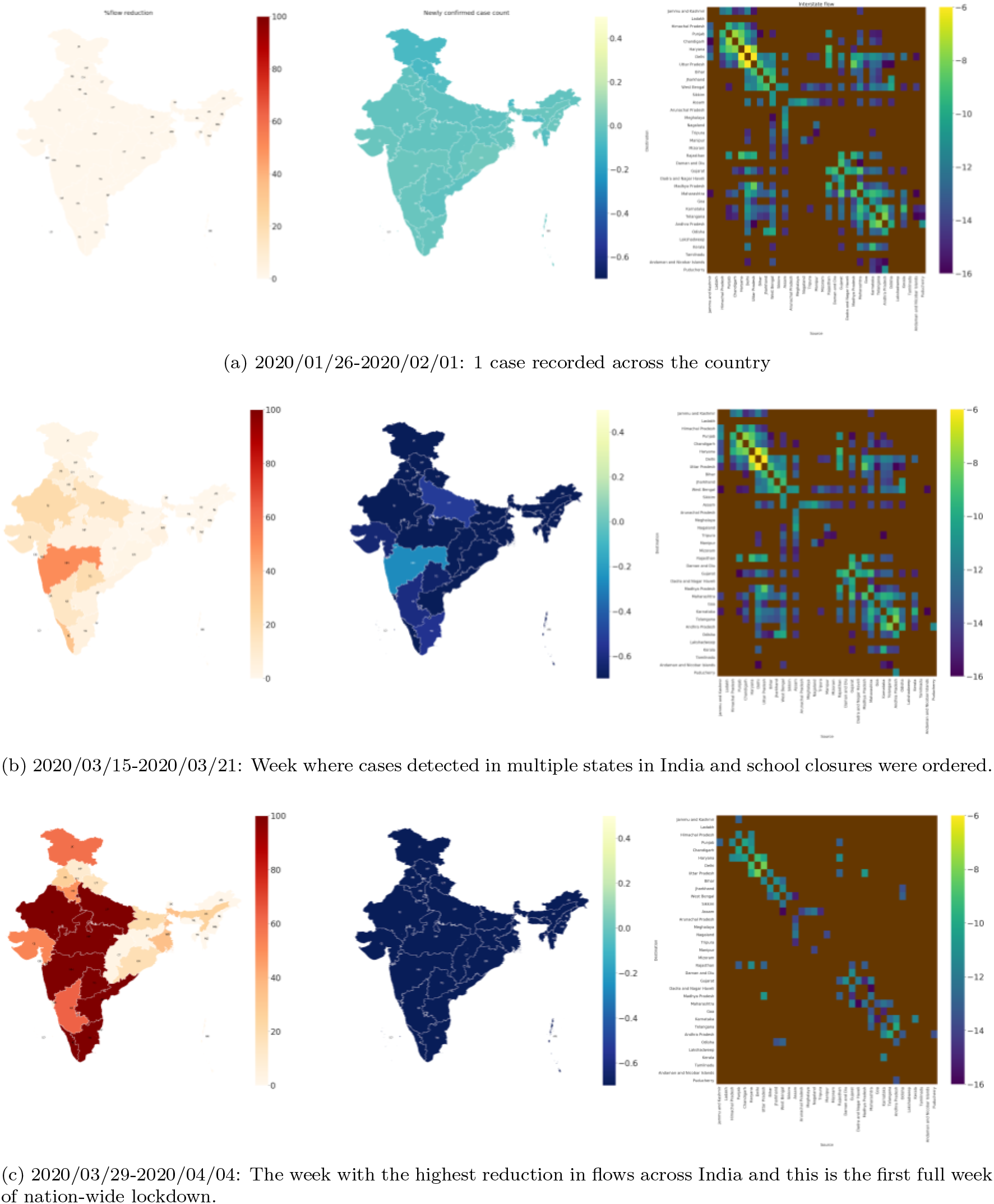
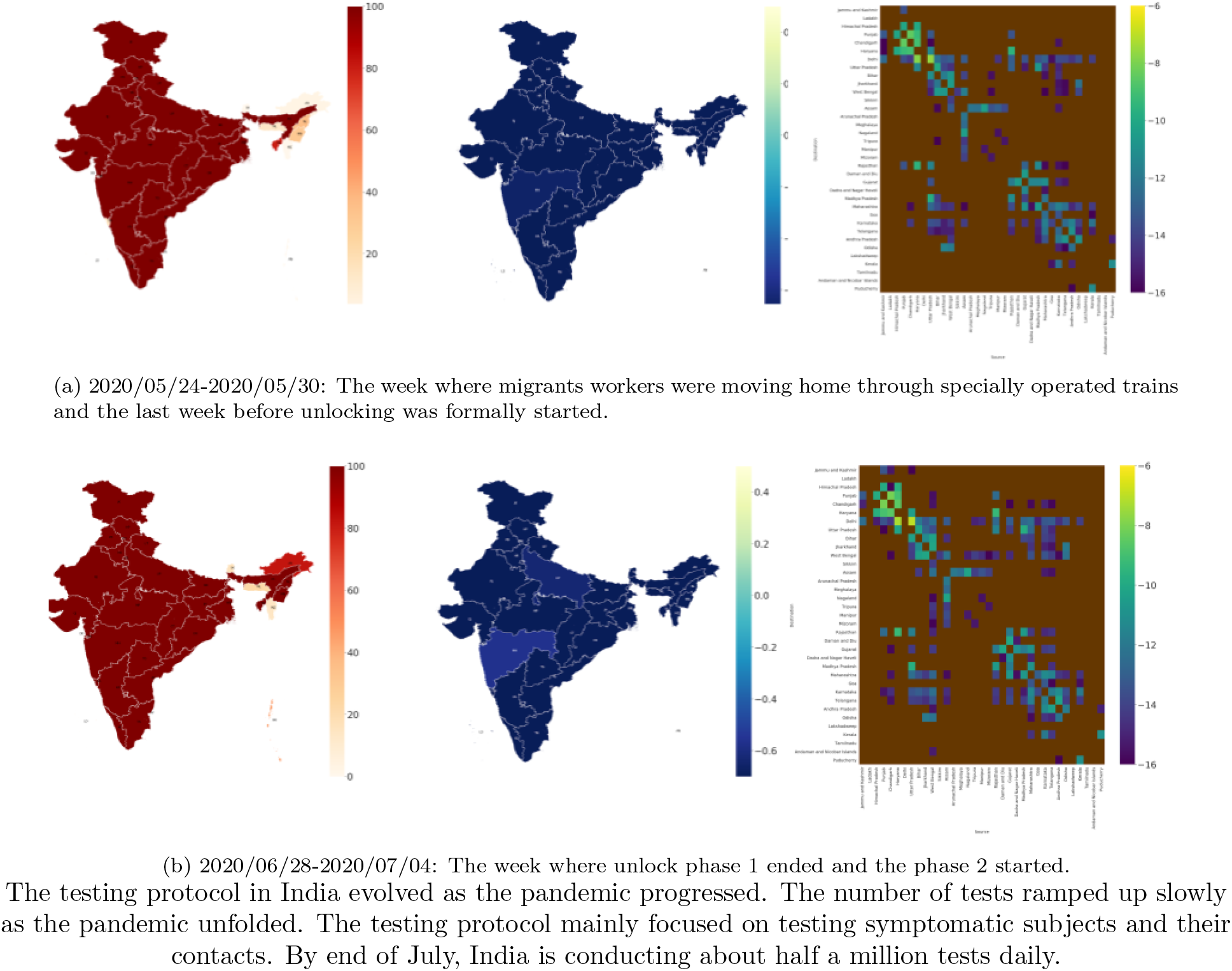
(**India Data**) A comparison of variation in number of new cases to variation in flow reduction and inter-state mobility across different weeks. Column 1 represents the spatial distribution of new cases across India, column2 shows the spatial distribution of FRR, and column 3 depicts the heatmap of natural logarithm of the flow between states (states are ordered in order to preserve a sense of adjacency as far as possible with self-loops suppressed and brown shade indicates state-pairs not recorded in the data.). Row 1 represents normal times (only a few cases recorded). Row 2 represents a week where cases started picking up in India along with reduction in mobility. By the week of 2020/03/29-2020/04/04, the weekly new cases has increased substantially and the nation-wide lockdown has brought down mobility substantially, with the numbers of state pairs with non-zero flows dropping by nearly 70% like in the US. As India starts special trains to take migrants back home, we observe increased mobility between certain states in the week of 2020/05/24-2020/05/30 (Row 4). The row 5 shows further increase in mobility between state pairs as India begins phase-wise unlocking.

## Footnotes

1 Source: https://covid19.who.int/ as of July 27, 2020.

2 https://www.un.org/development/desa/en/news/policy/wesp-mid-2020-report.html

3 Detailed global cases data available at https://nssac.bii.virginia.edu/covid-19/dashboard/

4 https://nssac.bii.virginia.edu/covid-19/dashboard/

5 We have selected a few illustrative countries to keep the discussion tractable, which were salient from an epidemiological perspective; additional country data can be reported if requested.

6 The starting dates of lockdowns in different countries were obtained from the https://en.wikipedia.org/wiki/COVID-19_pandemic_lockdowns

